# BMPR-II biomarkers for testing therapeutic efficacy in pulmonary arterial hypertension –the StratosPHere 1 study

**DOI:** 10.64898/2025.12.19.25342649

**Authors:** Rowena J. Jones, Eckart M.D.D. De Bie, Anthony Y.K.C. Ng, Benjamin J. Dunmore, Nina Deliu, Stefan Graf, Allan Lawrie, Joseph Newman, Gary Polwarth, Christopher J. Rhodes, Martin R. Wilkins, Anna Hemnes, James West, Sofia S. Villar, Paul D. Upton, UK National Cohort Study of Idiopathic and Heritable PAH Consortium, the Uniphy Clinical Trials Network, Mark R. Toshner

## Abstract

Pulmonary arterial hypertension (PAH) is a rare, life-limiting disease where deficiency of the TGF/BMP pathways have causal roles in hereditary and idiopathic forms. It is an attractive candidate for therapeutic intervention but there is an unmet need for clinically-relevant and practical biomarkers that can measure target engagement. A major challenge has been the inaccessibility of lung tissue in disease for molecular profiling. Here we explore the surrogate capacity of peripheral blood BMP pathway-specific markers. We demonstrate that BMPR-II in flow cytometrically characterised white blood cell subsets is reduced in a proportion of patients, however proteomic analysis demonstrates pleiotropic alterations of TGFβ/BMP modulators. Downstream BMPR-II canonical and non-canonical signalling is impacted and measurable in whole blood. We present discovery and international replication cohorts for a transcriptomic BMPR-II signalling signature. The composite biomarker panel is repeatable, reproducible, longitudinally stable and expressed in correlated gene modules in PAH which associate with clinical outcomes. The assay performance characteristics of the biomarker panel make it feasible for early phase, target engagement clinical trials and an adaptive three arm study of two pre- clinically validated modulators of BMPR-II is underway.

**One Sentence Summary:** The first demonstration of clinically-relevant BMP biomarker panels validated in international populations of patients with PAH.

## INTRODUCTION

Pulmonary arterial hypertension (PAH) is a rare progressive disease where patients rely on life-long treatment, often culminating in lung transplant or premature death. Cumulative pulmonary vascular endothelial dysfunction and remodelling causes obstruction of arterioles, placing significant burden upon the right heart, often resulting in heart failure. Whilst the genetic and molecular causes of PAH are increasingly well characterised, the translation to therapies that directly target underlying causes of disease is lagging. Standard of care, until very recently, remained with combinations of short-acting vasodilators with no demonstrable disease modifying benefit (*1*).

Imbalances in the transforming growth factor beta (TGFβ) superfamily receptor pathways; bone morphogenetic protein type-II receptor (BMPR-II) and activin type-II receptor (ActR-II) have been identified as prominent drivers in pulmonary vascular homeostasis and remodelling. Acting as inter-related pathways, circulating BMP and activin ligands exert their signalling via type-I and type-II transmembrane serine/threonine kinase receptors. Activation of the BMP pathway through BMPR-II, in complex with cognate type I receptors that define ligand selectivity, propagates signalling via phosphorylation of Smad1/5/8 and recruitment of the co-smad, Smad4, to regulate vascular homeostasis (*2*). This is balanced by activin and GDF ligands (GDF8 and 11), which bind activin receptor complexes to activate the Smad 2/3 – Smad4 pathway, promoting vascular cell proliferation (*3*). Loss-of-function mutations in the gene encoding the BMP type-II receptor (*BMPR2*) account for 53-86% of heritable PAH patients and occur in 14-35% of idiopathic cases (IPAH) (*4*),with further mutations occurring in the gene encoding activin-like kinase type-1 (ALK1) receptor (*ACVRL1*) (*5*), the co-receptor endoglin (*ENG*) (*6*), signalling intermediaries *SMAD1, SMAD4* and *SMAD9* (*7*) and ligand BMP9 ligand (*GDF2*) (*8*). Loss of function in the BMP signalling pathway and promiscuity of type-II receptors (*9*) along with increased activin type-II receptor ligands and reduction in the antagonist inhibin-α (*10*), cumulates in a proposed shift towards activin type-II receptor and Smad 2/3 mediated vascular remodelling. Recent trials targeting the TGFβ pathways through a first-in-class activin type-II receptor fusion protein ligand trap, sotatercept, has shown impressive clinical efficacy through to regulatory phase clinical trials (*11*). Pre-clinical data demonstrate reduced activation of the Activin A mediated Smad 2/3 pathway, with proposed rebalancing in favour of the anti-proliferative BMPR-II/BMP signalling pathway (*3*). However, these mechanisms have not yet been evaluated and proven in human clinical studies.

A major limiting factor has been the lack of target engagement assays for the TGFβ superfamily. It is neither safe nor ethical to sample the lung by biopsy in PAH and there is no molecular imaging developed to attempt non-invasive assessment. To date no alternative biomarker strategy has been validated. The emergence of potentially off-target angiogenetic and haematological side effects with sotatercept highlight the importance of a better understanding of TGFβ/BMP pathway engagement as we further develop therapies targeting these pathways. In light of this, the identification of mechanistic response biomarkers of drug activity to the TGFβ/BMPR-II pathways is required to refine the evaluation of future therapeutics. These will be critical in proving drug effectiveness, stratification of patients to the most relevant intervention for their treatment, and for better understanding of off-target adverse effects.

To address this need, the StratosPHere 1 study (NCT05767918) was set up as a precursor to proposed clinical trials treating PAH patients harbouring a *BMPR2* mutation with therapeutics preclinically demonstrated to increase cell surface BMPR-II; hydroxychloroquine (*12, 13*) and 4-phenylbutyrate (*14*), in a Bayesian response adaptive trial design. We report the development of peripheral blood biomarkers of BMP signalling and subsequently demonstrate a transcriptomic biomarker panel found to be dysregulated in two international prospective cohorts. Unsupervised machine learning using data from the UK’s National Cohort Study of Idiopathic and Heritable PAH (UK PAH cohort study) has enabled the stratification of patients based upon co-regulated gene modules in our biomarker panel which demonstrated associations with long term survival and clinical outcomes. As a direct readout, we also show detection of cell surface BMPR-II protein measured using spectral flow cytometry.

## RESULTS

### Identification of a transcriptomic biomarker panel of BMPR-II biology using quantitative PCR

Investigation of publicly available transcriptomic data indicates *BMPR2* and relevant type I co-receptors are well expressed in leukocyte populations: with ubiquitous expression of *BMPR2* and type I receptor *ACVR1* (ALK2), and high expression of type I receptor *ACVRL1* (ALK1) seen selectively in monocytes (Fig. S1A). To assess for the potential to detect BMPR-II pathway engagement we first determined if genes known to be downstream of BMPR-II pathway activation could be detected in peripheral blood samples for the purpose of building a biomarker panel using quantitative PCR (qPCR, Fig. 1A). We identified candidate genes of interest (GOI’s) from both relevant literature and from our previously published microarray study (*15*) (Fig. S1B).

**Fig. 1.**
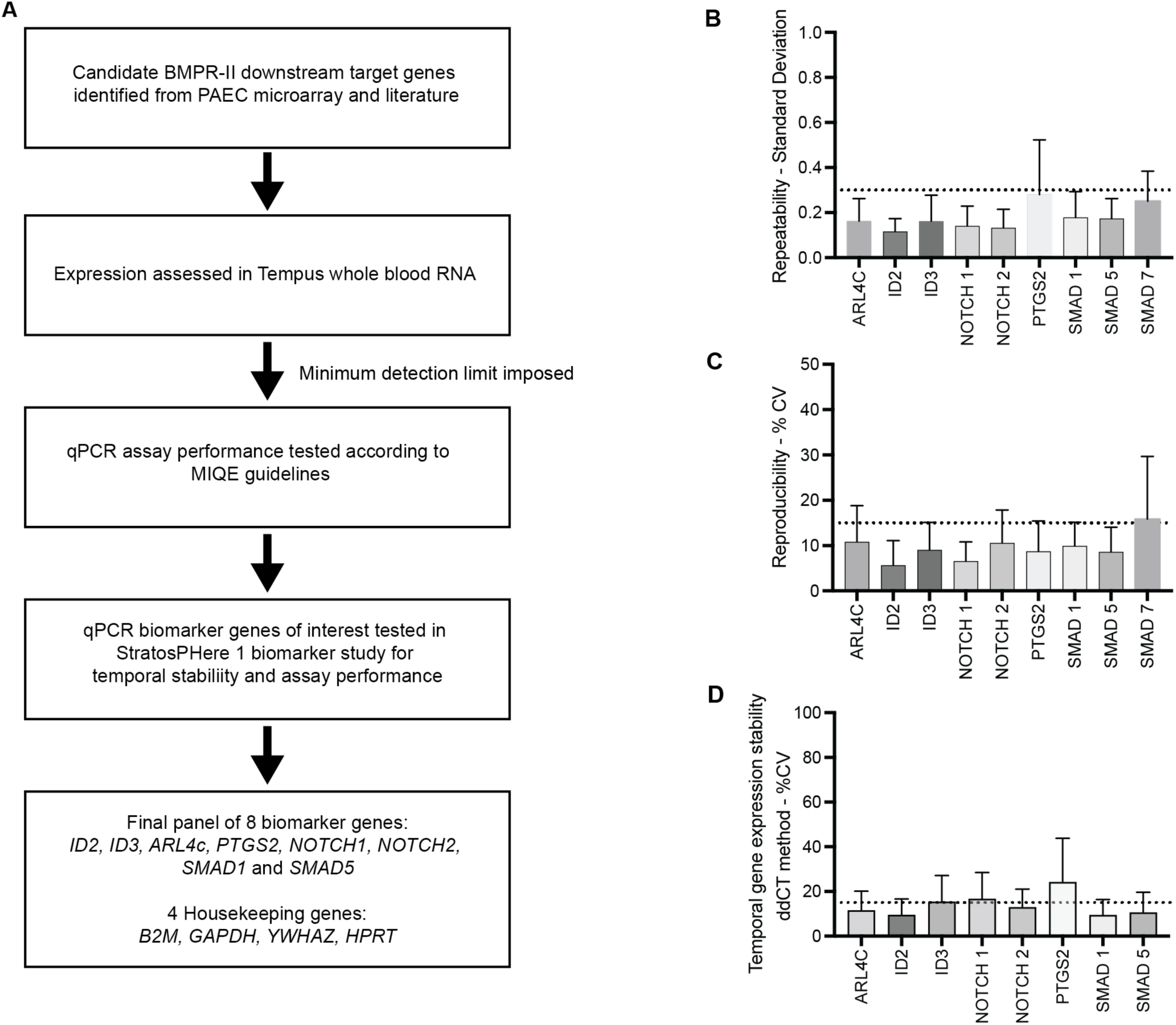
Development of a qPCR Biomarker Panel for the detection of BMPR-II target engagement in whole blood. **(A**) Workflow for the identification and development of qPCR biomarker panel for the detection of downstream engagement of the BMPR-II pathway. The candidate qPCR biomarker panel was tested in StratosPHere 1 samples (healthy controls N=15, and PAH patients N=20). Samples were assessed for (**B**) repeatability of technical replicates (standard deviation of N=4 replicates per sample, N=140 samples tested), with a cut-off of maximal 0.3 standard deviation for sample precision (dotted line) and for (**C**) reproducibility assayed across multiple runs (minimum N=8 samples assayed on two occasions) shown as co-efficient of variation (%CV) with a cut-off of maximal 15%CV (dotted line). (**D**) Temporal stability was assessed in PAH samples only (N=20) between baseline and 16 weeks (± 4 weeks) and shown as % CV dotted line at 15% CV.

For initial screening, expression of GOI’s was first assessed in peripheral whole blood RNA (N=6, Fig. S1C). GOIs with a readily detectable cycle threshold (Ct) value ≤30 were selected for further assessment. A comparison with RNA isolated from fractionated peripheral blood mononuclear cells (PBMCs) showed no enrichment of GOI’s compared to whole blood validating its use as a more practical clinical sample (Fig. S1C). Of the initial 22 candidate GOIs, 11 were readily detectable in whole blood RNA. Candidate GOIs were assessed for assay performance in StratosPHere 1 samples (for demographics see Table 1) according to MIQE guidelines (*16*). Technical replicability was assessed by determining the standard deviation (SD) of Ct values using a pre-defined maximal of 0.3SD (Fig. 1B). *EML6* and *SPRY1* were excluded at this point as Ct values exceeded 30 in the StratosPHere 1 cohort. Reproducibility of sample readouts was assessed for precision over repeated runs of qPCR and was tested in a minimum of 8 samples per gene with a pre-defined percentage coefficient of variation (%CV) maximum of 15% between repeated measures, at which point *SMAD7* was excluded (Fig. 1C).

**Table 1.** Demographics and clinical characteristics of participants of the StratosPHere 1 Biomarker study. Data shown as mean (range) or mean ±SD. Abbreviations: 6MWD – six-minute walk distance; mPAP – mean pulmonary arterial pressure; mRAP – mean right atrial pressure; PVR – pulmonary vascular resistance; NT-proBNP – N-terminal pro b-type natriuretic peptide; PDE5 inhibitor – phosphodiesterase 5 inhibitor; PRA – prostaglandin receptor antagonist.

|  | Healthy | PAH |
| --- | --- | --- |
| Number of subjects | 18 | 28 |
| Age at sampling (years) | 40 (26-58) | 53 (20-80) |
| Male:Female | 6:12 | 8:20 |
| Disease pathology |  | 8 *BMPR2* mutation causing-PAH<br>20 IPAH |
| WHO classification I/II/III |  | 7/6/15 |
| 6MWD (m) | | 449 ± 142<br>(Missing 4) |
| **Haemodynamics** |  |  |
| mPAP (mmHg) | | 46 ± 13<br>(Missing 7) |
| mRAP (mmHg) | | 6.95 ± 4.8<br>(Missing 7) |
| PVR (dynes/sec/cm<sup>-5</sup>) | | 624±312<br>(Missing 7) |
| NT-proBNP (pg/ml) | | 628±884 |
| **PAH targeted treatment** |  |  |
| PDE5 inhibitor |  | 12 |
| Endothelin receptor antagonists |  | 10 |
| Prostaglandins/ PRA's |  | 7 |
| Calcium channel blocker |  | 1 |
| Diuretic |  | 5 |
| Mono/dual/triple therapy |  | 13/7/1 |

The selected 8 candidate BMPR-II biomarker genes (*ARL4C, ID2, ID3, NOTCH1, NOTCH2, PTGS2, SMAD1* and *SMAD5*) were assessed for temporal stability between baseline and +16 weeks in the StratosPHere 1 PAH population (N=20) between time points calculated using both the ΔΔCT method and using a standard curve method (Fig. 1D-E, Fig. S1D-F, Table S1). Temporal stability of the PAH population was good in all genes tested with maximal %CV in *PTGS2* expression between time points (22.5%), but lower in other genes (range 9.4-16.7 %CV, ΔΔCT method). Pre-planned power calculations confirmed the validity of a composite multi-biomarker panel approach for our subsequent three-arm adaptive clinical trial (*17*). Little difference was observed when reporting variation using either the qPCR standard curve or ΔΔCT method, indicating validity in both normalisation methods. Long term stability was further validated using RNAseq data from both the UK PAH Cohort study (*18*) and the TRANSFORM trial (*19*) and no significant differences were observed between time points (Fig. S2).

### The BMPR-II biomarker panel is altered in a large validation cohort with distinct gene modules showing correlated expression patterns

We compared expression of our BMPR-II biomarker panel genes in a large cohort of PAH patients (N=356) with healthy controls (N=67) using RNAseq data from the UK PAH cohort study (*18*) (demographics Table S2). This revealed differential expression of 6 of the 8 GOIs (*ARL4C, ID3, NOTCH1, NOTCH2, PTGS2* and *SMAD5*, Fig. 2A).

**Fig. 2.**
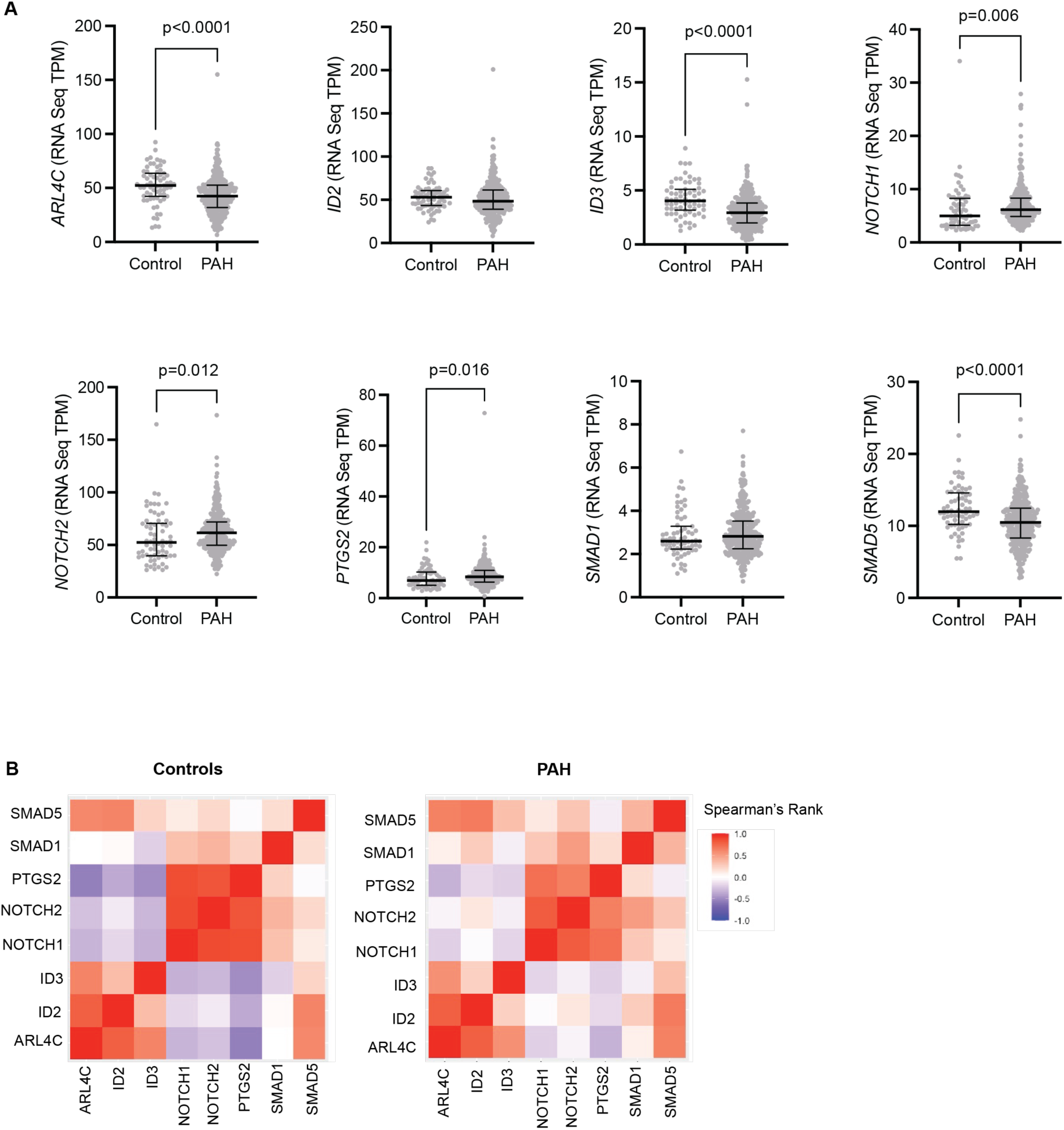
Differential expression of BMPR-II biomarker panel genes reveals gene modules. **(A**) Levels of BMPR-II biomarker panel genes measured in the UK PAH cohort study using RNAseq in controls (N=67), and PAH (N=356). Scatter plots show median ± IQR, statistical test Kruskal-Wallis. (**B**) Heat map of Spearman’s rank correlation of BMPR-II biomarker panel genes in controls (N=67) and PAH (N=356), where red shows positive correlation and blue shows negative correlation.

Spearman’s rank correlation identified significant correlations of gene expression in all cohort of patients and controls (Fig. 2B and Table S3); a positive correlation of *ID2, ID3* and *ARL4C* was highly distinct from a second positively correlated gene module of *NOTCH1, NOTCH2* and *PTGS2* (Spearman’s rho >0.8 in healthy controls). In control samples, these two correlated gene modules were highly negatively correlated with each other (mean Spearman’s rho across 9 combinations, −0.32), however, in PAH this decreased in significance with the dysregulated expression of the genes (mean Spearman’s rho across 9 combinations, −0.12). Of notable interest, novel gene *ARL4C* correlated strongly with known BMPR-II signalling genes; *ARL4C* with *ID2* (Spearman’s rho = >0.79, p=<0.0001) and *ID3* (Spearman’s rho = >0.56, p=<0.0001 in PAH), and *PTGS2* correlated with *NOTCH1* and *NOTCH2* (Spearman’s rho = >0.71, Spearman’s rho = >0.64 and p=<0.0001 respectively).

### Unsupervised clustering of our BMPR-II biomarker panel genes reveals distinct subgroups in PAH

Utilising the RNAseq data from the UK PAH cohort study we performed unsupervised machine learning to cluster patients based upon their transcriptomic expression of our BMPR-II biomarker panel genes. After comparing multiple clustering algorithms using Euclidean distances, Kmeans clustering with three clusters was identified as superior to partitioning around medoids and hierarchical clustering (Fig. S3A-F) and was applied to PAH patient data (N=356). Patients were found to cluster based upon distinct patterns of biomarker gene expression. When compared between clusters, patients in cluster 1 (N=145) had highest levels of *NOTCH1*, *NOTCH2*, *PTGS2* and *SMAD1* expression (p <0.0001) (Fig. 3A, Table S4) and patients in cluster 3 (N=72) had lowest the expression of *ID2*, *ID3*, *ARL4C* and *SMAD5* (p <0.0001). When compared to healthy control biomarker gene expression cluster 2 (N=138) had the most similar expression profile (Fig. S3G,H). Gene changes driving formation of the patient clusters appeared to coincide strongly with the gene modules observed in our correlation analysis, suggesting that orchestrated co-expression of these gene modules is important for disease pathology.

**Fig. 3.**
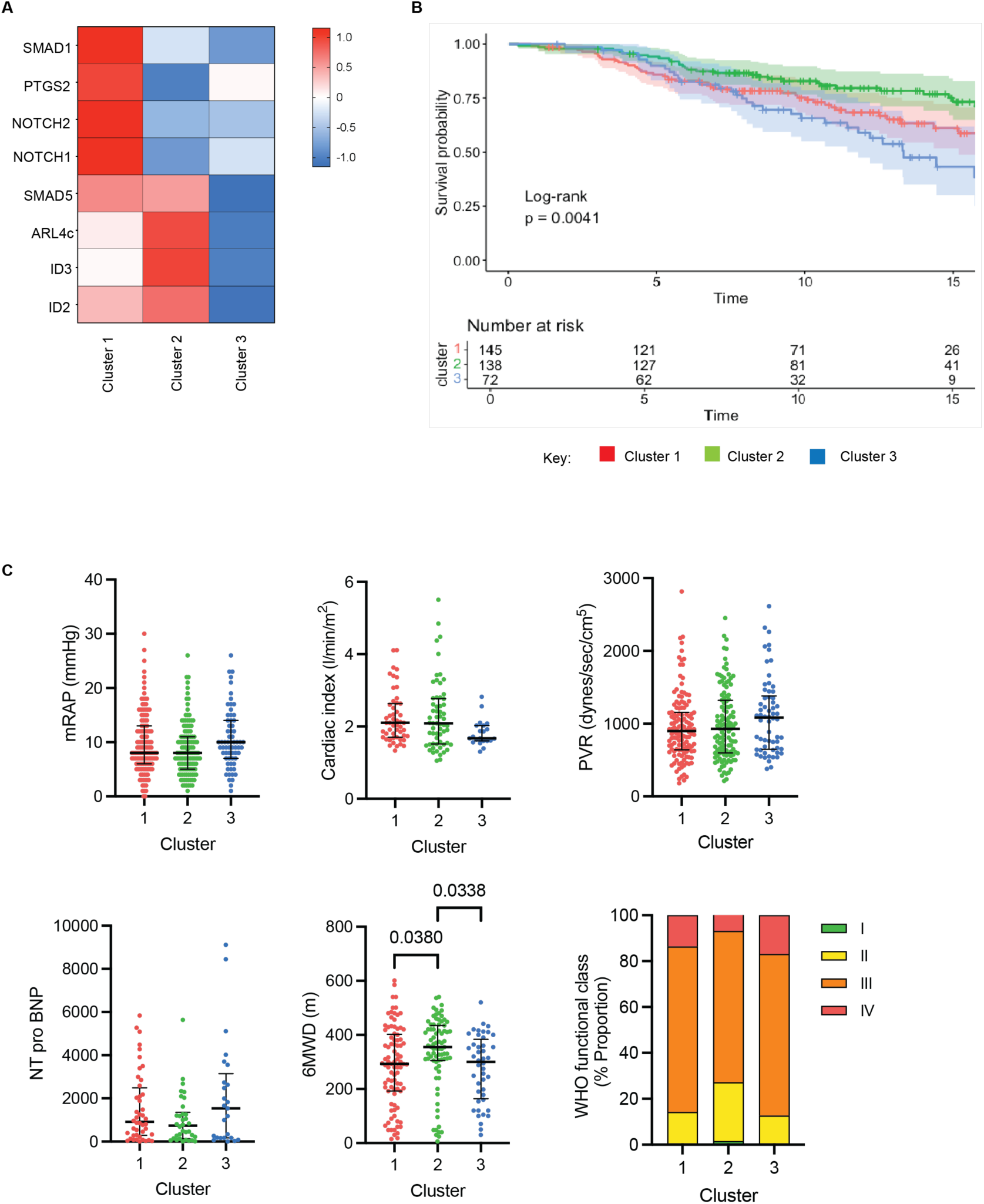
BMPR-II biomarker panel genes cluster into distinct groups based upon expression profile with significant impact on survival and haemodynamic outcomes. Unsupervised clustering performed on biomarker panel genes using RNAseq data from the UK PAH cohort study (N=356). **(A)** Heatmap of Log10 gene expression compared between the three clusters using Z-score. (**B)** Kaplan-Meier survival analysis for patients stratified by cluster. Statistical analysis was performed using pairwise log-rank test. **(C)** Haemodynamic and functional characteristics between patient clusters: right atrial pressure (mRAP), cardiac index, pulmonary vascular resistance (PVR), NT pro-BNP, 6-minute walk distance (6MWD) and WHO functional class. Data shown as median ± IQR or % proportion. Kruskal-Wallis statistical test used with Dunn’s multiple comparison test where appropriate. Chi squared test shown for functional class.

*BMPR2* transcript expression and the number of BMPR-II mutation carriers also varied between clusters. Whilst *BMPR2* transcript levels do not differ significantly between controls and PAH (Fig. S3I), patients in cluster 3 were found to have significantly reduced *BMPR2* (Fig. S3J) and the highest proportion of *BMPR2* mutation carriers (27.8% vs. 16.4% and 19.6%, cluster 3 vs clusters 1 and 2, Table S5).

### Clustering of patients based upon BMPR-II biomarker genes shows significantly differing clinical characteristics and survival outcomes

We next assessed the relationship between patient clusters with mortality and clinical characteristics (Fig.3, Table S5). Kaplan Meier survival curves demonstrated significantly different survival for patients between clusters (Log rank = 0.0041, Fig. 3B). Poorest survival was observed in patients in cluster 3, with a survival at < 50% after 15 years. Patients in cluster 2 had the best survival at > 75% after 15 years. Hemodynamic characteristics were not statistically different, though with a consistent direction of effect in cluster 3 for right atrial pressure (mRAP), cardiac index, pulmonary vascular resistance (PVR) and NT-proBNP (Fig. 3C, Table S5). Patient 6-minute walk distance and functional class differed significantly between clusters of patients, with patients in clusters 1 and 3 demonstrating significantly shorter walk distance (cluster 1 median 293m; cluster 2, 355m; cluster 3, 300m, Fig. 3C). Similarly, a higher proportion of WHO class III and IV were also observed in clusters 1 and 3 compared to cluster 2 (Fig. 3C). Cox regression modelling confirmed the poorer prognosis for patients in cluster 3 after correction for age at diagnosis and sex (odds ratio 2.2, 95% confidence interval 1.38-3.6, p<0.001). Age at diagnosis remained a significant factor in early mortality (p <0.001) (Fig. S3K). Internal validation of 1000-repeat random sampling of the clustered patients confirmed overall differences in survival probability between clusters, with robustly poorer survival in cluster 3 (p= <0.0001) (Fig. S3L).

### Validation of dysregulation of our BMPR-II biomarker panel in an independent cohort

Our novel BMPR-II biomarker panel was validated using qPCR in an external cohort of 38 group 1 PAH patients and 16 healthy controls (Table S6). The validation cohort had a broader disease aetiology compared to the UK PAH cohort study, and comprised 51% IPAH/HPAH, with a further 43% diagnosed with connective tissue disease (CTD) PAH. Significant and more pronounced differential expression was observed for *ARL4C, ID2, ID3, PTGS2* and *NOTCH1*. As previously observed, there was no statistically significant difference in *SMAD1* expression. No statistically significant difference was recapitulated for *NOTCH2* or *SMAD5* (Fig. S4).

### Detection of cell surface BMPR-II protein in leukocyte cell populations reveals high expression in myeloid and B-cell populations that is significantly reduced in PAH which show disease sensitive clustering of expression and adequate technical performance for biomarker development

We utilised spectral flow cytometry to identify classes of leukocyte subsets from PBMC fractions and assessed cell surface BMPR-II expression using a monoclonal antibody to the BMPR-II extracellular domain (ECD) in StratosPHere 1 samples. Cell surface staining revealed BMRP-II was highest in CD14^+^ CD16^-^ classical monocytes, with 100% positive staining (Fig. 4A, representative histograms). Levels were also high in CD11C^+^ dendritic cells, CD14^-^ CD16^+^ non-classical monocytes and CD19^+^ B-cells where staining profiles exhibited a partial / biphasic staining pattern. Staining was lowest in CD3^+^ CD56^+^ natural killer T cells, CD56+ CD16^+^ natural killer cells and CD3^+^ T cells (Fig. S5A).

**Fig. 4.**
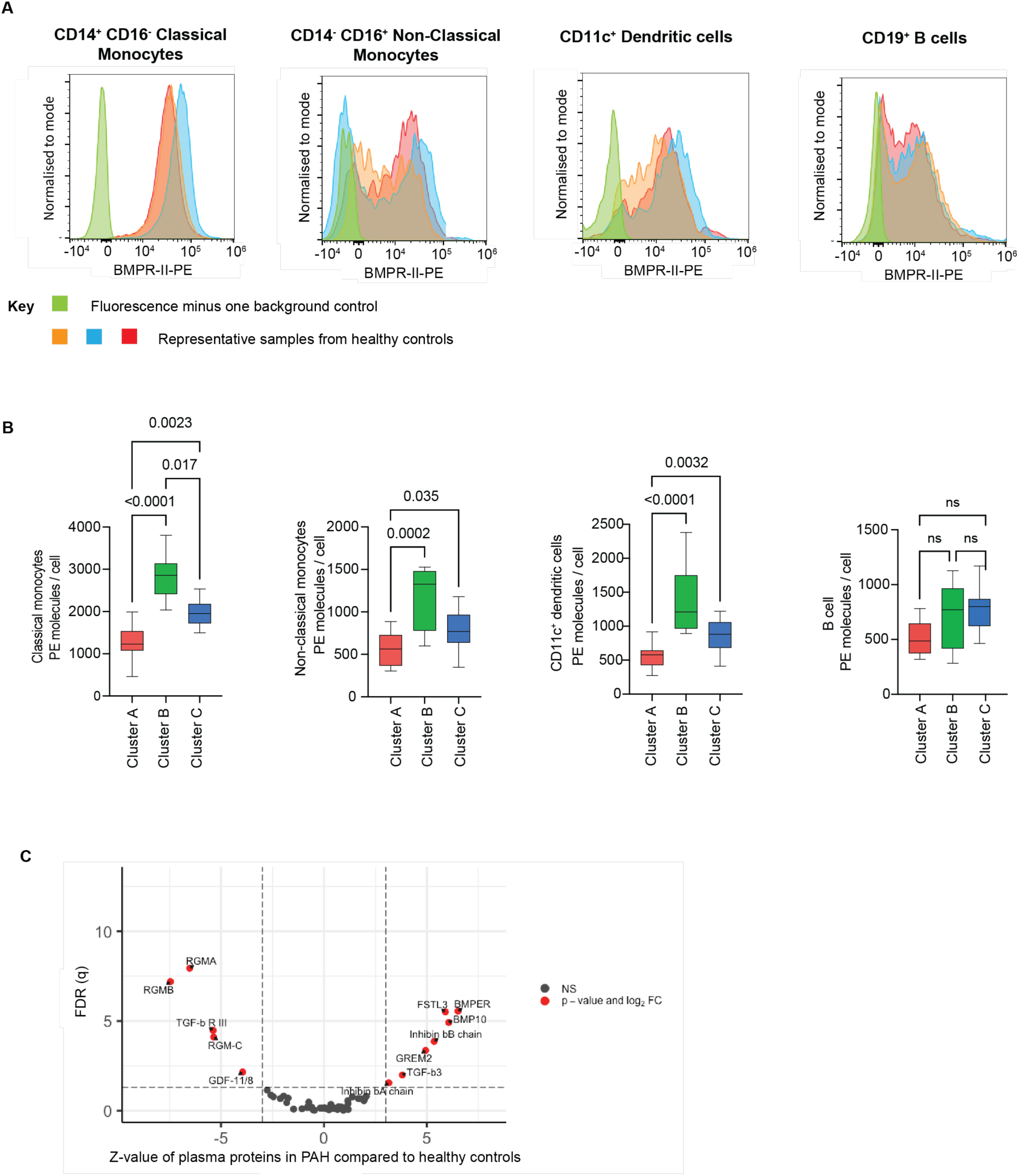
Cell surface BMPR-II is detectable in major leukocyte subsets and is reduced in PAH and the TGFβ plasma proteome is highly dysregulated. Immunophenotyping of peripheral blood mononuclear cells was used to determine levels of cell surface BMPR-II in CD14^+^ CD16^-^ classical monocytes, CD14^-^ CD16^+^ non classical monocytes, CD14^-^ CD11c^+^ dendritic cells and CD19^+^ B-cells. **(A)** Histogram plots demonstrate BMPR-II-PE fluorescence signal of 3 example healthy controls (blue, orange and red peaks) versus a fluorescence minus one gating control (green peak). (**B)** Cell surface levels of BMPR-II compared between clusters generated from unsupervised machine learning of combined healthy controls (N=17) and PAH patients (N=28) in classical monocytes, non-classical monocytes, dendritic cells and B-cells. BMPR-II is represented as PE molecules per cell of the positive gated population taken at one time point, median ± IQR. Statistical analysis performed using Kruskal-Wallis test and Dunn multiple comparisons test. (**C)** Volcano plot of the Z-score change in PAH (N=357) standardised to healthy controls (x axis) versus false discovery rate (FDR; y axis) of data derived from plasma proteome array in the UK PAH cohort study (*23*). Dotted lines are drawn at Z-score ± 3 and FDR q<0.05 and proteins denoted in red exceed these thresholds.

Correction for BMPR-II antibody batch was performed, and cell surface BMPR-II levels were compared between controls (N=17) and PAH patients with and without mutations in *BMPR2* (N=20 / N=8 respectively) in the cell populations highly expressing BMPR-II. Cell population specific expression was not significantly decreased in patients (Fig. S5B). *BMPR2* mutations have been found to affect cell surface localisation in different ways depending upon the location of the mutation (*20, 21*) and decreases in lung BMPR-II expression are identified in non-mutation carriers (*22*). To address this, clustering analysis of the antibody batch corrected BMPR-II cell surface staining residuals from monocytes, dendritic and B-cells of patients and healthy controls was performed. After comparing multiple clustering algorithm permutations using Euclidean distances, Kmeans clustering with three clusters was identified as superior (Fig. S5C-H), generating 3 distinct clusters (A-C).

We observed statistically significant differences in cell surface BMPR-II staining between clusters in myeloid lineage cell subsets: classical monocytes, non-classical monocytes and dendritic cells (Fig. 4B, Table S7). The clustering analyses demonstrated that individuals in cluster A had significantly reduced levels of cell surface BMPR-II compared with individuals in clusters B and C. A similar directionality was observed in B-cells (p=0.085). Comparison of patient and control distributions between clusters showed cluster A to be enriched with PAH patients (13 out of 15), compared with clusters B and C. Of the 28 patients analysed, 45% of IPAH and 50% of the *BMPR2* mutation carriers demonstrated significantly reduced cell surface BMPR-II and grouped in cluster A. We noted a high proportion of *BMPR2* mutation carriers with N terminus mutations (W9X, C123R, R321X, R332X) in cluster A, compared with clusters B and C which had higher cell surface BMPR-II (Fig. S5I). To characterise the BMP and BMPR-II signalling pathway in blood derived cells we utilised the monocytic-like THP1 cell line to assess the effect of BMP ligand stimulation on the cell lineage with highest BMPR-II expression. Transcriptomic TGFβ receptor expression in THP1 cells demonstrated highest levels of *BMPR2* and *ACVRL1* and *ACVR1* (Fig. S1A). Based upon the profile of type I and II receptors we confirmed ligand responses to BMP6 and BMP9 in the majority of the BMPR-II biomarker panel (Fig. S6).

To clarify the potential of flow-cytometric cell surface BMPR-II measurement to complement our functional pathway activation biomarker panel, we determined assay repeatability and temporal stability. A pre-defined technical variability threshold of 15%CV was passed by all high expressing cell types (B-cells, dendritic cells and classical and non-classical monocytes and dendritic cells, N=69, Fig. S5J). In paired baseline to week 16 samples temporal stability was also acceptable at less that 15% CV in B-cells and classical and non-classical monocytes in patients, but above threshold in dendritic cells (16.7%CV) (Fig. S5K, N=15).

### Circulating proteins associated with PAH highlights dysregulation in control of the BMP / Activin pathways

Whilst we have described BMPR-II relevant biomarkers relating to cell surface protein expression and pathway activation it is pertinent to consider the microenvironment within the plasma in PAH that may further impact upon signalling pathways. Utilising findings from a recent study of an aptamer based plasma proteomic screen in patients from the UK PAH cohort study (N=357) (*23*) we examined proteins specifically linked to the TGFβ pathway. Of 63 proteins detected, 14 showed significant dysregulation in PAH (FDR q<0.05, Fig. 5, Table S8). Notably, members of the repulsive guidance molecule family (RGM), key co-receptors that enhance BMP signalling (*24–26*), are down regulated in PAH (RGM-A z-value −6.525, FDR q=1.14×10^-08^; RGM-B z-value − 7.459, FDR q=6.36×10^-08^; RGM-C z-value −5.357, q=FDR 7.91×10^-05^). Comparatively, BMP modulators, BMPER (*27*) and gremlin-2 (*28*) are increased (z-value 6.512, q=FDR 2.81×10^06^ and z-value 4.939, q=FDR 0.00044 respectively).

These findings may indicate that in PAH there is a larger picture of TGFβ pathway dysregulation that exists on genetic, transcriptomic and proteomic levels. It is currently unknown if the presence of dysregulated pathway regulators is cause or consequence of disruption to the BMPR-II pathway, or whether they may be resolved with restoration of BMPR-II. This strongly suggests that measurement of BMPR-II protein alone may be insufficient for proving therapeutic efficacy, and the requirement of a multi-approach biomarker to measure downstream pathway target engagement or resolution, such as our qPCR biomarker panel will be more informative.

## DISCUSSION

A major challenge of the translation of preclinically validated therapeutics into clinical trials is the development of practical biomarkers that demonstrate proof of mechanism and are suitable for use as early phase trial endpoints. In this study we have generated a comprehensive, easily measured, peripheral blood target engagement panel for BMP signalling that is repeatable, reproducible and longitudinally stable. We have demonstrated for the first time that BMP signalling in peripheral blood is abnormal at gene transcript, cell surface and plasma proteome levels across patients with group 1 PAH. Moreover, discrete gene modules from our transcriptomic biomarker panel are co-regulated across health and disease and are informative of patient outcomes. The targeting of the TGFβ superfamily/BMPR-II signalling axis is the most exciting current therapeutic development in PAH, potentially relevant across common and rare forms of the disease. The clinical relevance of this is clear with the first licensed therapy targeting the activin/BMP axis; sotatercept, and the use of our defined biomarkers is currently being tested in the StratosPHere 2 study (IRAS 1003631).

Sotatercept has become the first intervention to successfully target the activin/BMP signalling pathway with demonstrable clinical efficacy, showing significant improvements in 6-minute walk distance, PVR and NT-proBNP (*11*). *In vitro* models of sotatercept treatment demonstrate that the activin binding molecule sequesters circulating activin A/B and GDF8/11 (*3*). However, its binding profile potentially extends to other BMP molecules (*29*) and the appearance of telangiectasias and bleeding events is concerning, whereby sequestration of BMP9/BMP10 may inadvertently result in an hereditary haemorrhagic telangiectasia (HHT)–like phenotype. BMP9/10 are high affinity ligands of TGFβ type-I receptor, ALK1 and mutations in the gene encoding ALK1 (*ACVRL1*) are a common cause of HHT. Indeed, treatment with an ALK1-Fc ligand trap, dalantercept, in cancer patients, bound BMP9/10 and inhibited endogenous ALK1, but resulted in telangiectasias in 8 of 37 patients (*30*), a recognised on-target pharmacodynamic effect. Given the significant advances of sotatercept and the ongoing development of therapies with subtly different ligand binding profiles such as KER-012 (Keros Therapeutics), a better understanding of target engagement in humans is required to clearly identify the mechanism most relevant to PAH pathobiology. Whilst there is strong evidence for inhibiting the potentially pathogenic activin/GDF pathway, there is no human evidence for the rebalancing of BMPR-II expression of signalling and nor are the related side effects understood. Understanding on- and off-target effect on the relevant pathways will be critical to developing this therapy area in the future.

The StratosPHere 2 trial, a first in class design for PAH, will test the effect of two pre-clinically validated repurposed drugs; hydroxychloroquine (*12, 13*) and 4-phenylbutyrate (4PBA) (*14*) on their ability to increase cell surface BMPR-II signalling and cell surface expression in patients stratified by *BMPR2* mutation type using a Bayesian response adaptive methodology. Based upon our findings we will utilise our composite transcriptomic biomarker panel for readout of BMP/BMPR-II signalling measuring *ARL4C, ID2, ID3, NOTCH1, NOTCH2, PTGS2, SMAD1* and *SMAD5* gene expression as a primary endpoint in the study.

The application of unsupervised machine learning using RNAseq data revealed subgroups of patients with similar biomarker levels that corresponded to gene modules of similarly correlated genes. Low expression of *ID2/ID3* in combination with low *ARL4C*, *SMAD1* and *SMAD5* expression (cluster 3) was significantly associated with worsened clinical haemodynamics, shorter 6-minute walk distance, higher WHO functional class and demonstrably lower survival. *ID2* and *ID3*, members of the *Inhibitors of DNA binding family*, are commonly used as readouts for canonical BMPR-II signalling in vascular cells (*31*). Reduced *SMAD5* has been previously described in PAH (*18*), with levels linked to an eQTL SNP genotype and is biologically plausible as a readout of BMPR-II dysregulation as it encodes an intracellular transcription modulator that is activated as part of canonical BMPR-II signalling, along with *SMAD1*. Comparatively, *ARL4C* is a novel gene in PAH and was identified as being significantly reduced in PAECs treated with BMP9 (*32*). Interestingly, the highest proportion of *BMPR2* mutation carriers (27%), and lowest overall *BMPR2* transcript expression occurred in the reduced *ID2/ID3* cluster (cluster 3).

Patients in cluster 1 had highest levels of *BMPR2* transcript and proportionally fewer *BMPR2* mutation carriers and were characterised by raised *NOTCH1, NOTCH2, PTGS2* and *SMAD1* levels and were non-significantly associated with poorer survival and clinical outcomes compared with patients in cluster 2. *NOTCH2* has been shown to be increased in pulmonary arterial smooth muscle cells in the presence of BMP6, exacerbated by TNFα on a background on BMPR-II deficiency (*33*), and both *NOTCH1* and *NOTCH2* expression is increased in disease. NOTCH genes have been implicated in lung vascular remodelling in PAH and have been shown to functionally interact with BMPR-II for vascular endothelial cell regeneration (*34, 35*). *PTGS2* is the second novel gene in our panel to be linked to BMPR-II. Encoding cyclooxygenase 2, *PTGS2* has a key role in the conversion of arachidonic acid to prostaglandin E2, a key vasodilatory pathway affected in PAH and a pathway on which many common PAH drugs work. It is increased in response to BMP9 in PAECs (*32*), and is induced in PAECs, along with canonical BMPR-II target genes when treated with a BMPR-II signalling regulator compound, BUR1 (*36*). An unexpected link to vasoactivity in BMP signalling may require further work as currently BMP signalling is thought not to involve vasodilation. Expression of *PTGS2* was not found to be affected by PAH medications.

Comparatively, patients in cluster 2, accounting for approximately 40% of our cohort had biomarker levels most akin to healthy controls with the best physiological parameters and highest survival probability. This finding may be important for both the ontology of the disease as it may represent a non-BMP causal role in a subset of patients, but also in the stratification and selection of patients to future trials regarding the likelihood of detecting a response to treatment specifically targeting the BMP/BMPR2 pathway. The relevance of the observed gene modules may also be important in gauging treatment response. For example, the use of a composite panel of our chosen 8 biomarkers might not fit all patients, whereas observing change in the combination of *ID2/ID3/ARL4C/ SMAD5* module or the *NOTCH1/NOTCH2/SMAD1/PTGS2* module might be more informative depending upon the transcriptomic profile of patients prior to treatment.

Though we have demonstrated that direct assays of cell surface BMPR-II are feasible we question how informative they will be in translational research. The use of flow cytometry is known to be influenced by multiple technical factors, including antibody lot variation and daily variations in cytometer voltages. Detection of cell surface BMPR-II is inherently technically more complex than qPCR and so lends itself less to the use as a biomarker in clinical trials, despite showing acceptable levels of temporal stability. The changes in cell surface BMPR-II are subtle with enrichment of disease endotypes in the low expression clustering analyses. Focusing on BMPR-II expression but not downstream function is, in addition, likely to miss the effects of additional hits in the pathway, for example by many of the regulators changed in disease. The profiling of the TGFβ/BMP plasma proteome demonstrates the wider importance of pathway regulators and inhibitors in. Levels of RGMs, positive regulators of BMP signalling (*37*) were decreased, whilst levels of BMP antagonists BMPER (*38*) and gremlin-2 (*39*) were increased. Such imbalances may or may not be a consequence of direct BMPR-II pathway dysregulation, but their presence accords with the significant reduction in downstream gene expression despite modest overall reduction in TGFβ/BMPR-II itself.

Potential criticism of our work includes the diverse and different patient population in the validation cohort (which includes other group 1 causes of PAH), though we would argue that this broadens relevance to more than just idiopathic and heritable disease and has implications for therapy development. A limitation of our approach is that loss of BMPR-II signalling in disease is complex and still not completely understood in target tissues or otherwise. Although we demonstrate consistent findings that diverse peripheral blood biomarkers of BMPR-II pathway function are dysregulated, we do not yet understand the effect sizes and how they related to TGFβ/BMPR-II pathway focused treatments.

In summary, we report on peripheral blood-based biomarkers representative of BMPR-II pathway expression signalling and dysregulation in PAH at cell surface, proteome and gene expression levels. Gene expression correspond to clinical disease severity and survival. Our whole blood assessment of BMPR-II biology should allow for accessible measurement of therapeutic target engagement and offer an important validation of the importance of the TGFβ/BMP pathways in PAH.

## MATERIALS AND METHODS

### Study Design

The objective of this study was to identify and validate a biomarker or panel of biomarkers indicative of BMPR-II biology in PAH for use in clinical trials. To investigate this, the StratosPHere 1 biomarker study ran between April and December 2021 at the Royal Papworth Hospital, UK. A total of 35 PAH patients and 33 healthy controls were enrolled, including patients with idiopathic PAH (IPAH) and *BMPR2* mutation causing PAH. Of these, up to 28 PAH patients and 18 healthy controls were included in this study with both baseline (T0) and 16 weeks ± 4 weeks to simulate the duration of a clinical trial. Demographics and clinical characteristics are described in Table 1. All patients were deemed stable and treated with standard of care therapies. Peripheral blood samples were collected for Tempus whole blood RNA, isolation of peripheral blood mononuclear cells (PBMCs), plasma and serum. All samples were prepped within 4 hours of collection and samples were stored frozen and analysed in batches to minimise variation. Only samples where sample preparation had failed were excluded from further study. Preparation of blood samples is described in the supplementary methods.

For the initial testing of transcriptomic biomarker expression in whole blood N=6 healthy volunteers were analysed and RNA isolated from PBMC fraction and Tempus RNA whole blood were compared. A threshold of <Ct 30 was imposed. For the testing of readily detectable candidate biomarker stability in samples baseline and 16-week samples were prepared and run together to limit variation. For analysis of transcriptomic biomarkers using qPCR samples, we analysed samples over two batches with standard curves included for the purpose of normalisation. Individual samples were assayed in quadruplet and all samples for a given gene were analysed on the same plate. To test biomarker repeatability, reproducibility and temporal stability we determined standard deviation between quadruplet samples to test repeatability and imposed a 0.3SD threshold. Co-efficient of variation between repeat samples was used to test reproducibility with an imposed threshold of 15%, and temporal stability was compared between baseline and 16 weeks and described as %CV and fold change using both the ΔΔCt method and standard curve methods of qPCR analysis. Longitudinal stability was also confirmed using RNAseq values for genes of interest taken as part of the UK National cohort study of idiopathic and heritable PAH (www.ipahcohort.com) and the TRANSFORM study (*40*). Expression of transcriptomic biomarker genes was examined in monocytic cell line THP1 both at steady state and in the presence of BMP ligand stimulation using a minimum of 3 technical replicates.

Bioinformatic analysis was performed using whole blood RNAseq data collected from patients enrolled in the UK PAH cohort study (N=356 PAH including N=71 *BMPR2* PAH mutation carriers measured at single time point at or close to point of diagnosis) healthy controls (N=67). Analysis of expression of selected biomarker genes was compared between groups and unsupervised machine learning was used to identify patient clusters based upon expression of our selected BMPR-II biomarker genes. Patients stratified by cluster were examined for differences in survival and clinical characteristics. No patients were excluded from this analysis and where parameters were not available these are reported as missingness. Where patients were separated into a *BMPR2* mutation group, these patients had an identified genetic sequence mutation.

In addition to testing for a panel of genes representative of BMPR-II pathway activation, we also developed methodology for the detection of cell surface BMPR-II in leukocyte populations. PBMC samples collected in StratosPHere 1 were probed for surface BMPR-II levels (N=17 healthy controls and N=28 PAH) and where available longitudinal samples were compared to assess stability. Immunophenotyping was utilised to identify broad classes on leukocyte populations, only samples where yield was poor or immunophenotyping staining failed (N=2) were excluded. Post-hoc analysis of cell surface staining was performed using unsupervised machine learning to identify clusters of patients and healthy volunteers with differing levels of surface BMPR-II staining.

### Ethics

StratosPHere 1 samples were obtained following informed consent (clinicaltrials.gov NCT05767918). Samples data utilised from the UK National Cohort Study of Idiopathic and Heritable Pulmonary Arterial Hypertension were collected following informed consent (clinicaltrials.gov NCT01907295; UK REC Ref. 13/EE/0203) and/or the Sheffield Teaching Hospitals Observational Study of Pulmonary Hypertension, Cardiovascular and other Respiratory Disease (UK REC Ref 18/YH/0441). External validation of the BMPR-II biomarker panel was performed at Vanderbilt University Medical Centre, USA using patient samples collected with written ethical consent under the following projects (IRB #111530, #140983 and #170961).

### Measurement of transcriptomic biomarkers

RNA was reverse transcribed into cDNA using High-Capacity Reverse Transcriptase (ThermoFisher Scientific) according to manufacturer’s instructions. Multiple pooled RNA samples were included as a no-reverse transcriptase control to control for presence of genomic DNA contamination. cDNA samples were sub-aliquoted and stored at −20°C and repeated freeze thawing was minimised. Gene expression was determined by qPCR were performed using Power Up SYBR green master mix (ThermoFisher Scientific) with the following PCR temperature cycling: 50°C for 2 minutes, 95°C for 2 minutes; followed by 45 cycles of 95°C for 25 seconds, 55°C for 25 seconds, 72°C for 1 minute; followed by 95°C for 15 seconds, 60°C for 1 minute, 95°C for 15 seconds melt curve on a QuantStudio6 (ThermoFisher Scientific). Development of qPCR assays were designed to MIQE guidelines (supplementary methods). Sequences of primers are provided in (Table SM1). Cycle threshold (Ct) values were recorded. Normalisation of genes of interest was performed against the average Ct of four housekeeping genes (*B2M, GAPDH, HPRT* and *YWHAZ*), or two housekeeping genes for *in vitro* studies (*B2M* and *GAPDH*). Development of housekeeping genes for use in whole blood assays and relative gene expression performed using either the ΔΔCt method or the standard curve method is described in the supplementary methods.

### RNAseq data processing and unsupervised machine learning

R/R-studio was used for statistical analysis. RNAseq data had been previously pre-processed (*18, 41*). Transcript per million (TPM) data for BMPR-II biomarker panel genes in 356 PAH patients were used to perform unsupervised machine learning clustering. TPM values were first transformed by log–scaling. The ‘factoextra’ (v1.0.7) R-package was used to generate silhouette and elbow plots to test three clustering algorithms (partitioning around medoids, hierarchical clustering and K means) followed by identification of optimal clustering algorithm using the R package ‘ConsenusClusterPlus’ (v1.66.0), with the following parameters; maxK=6, reps = 1000, pItem =0.8 with Euclidean distances. Kmeans with K=3 was found to be optimal (Fig. S3A-E). Survival analysis was performed using R package ‘Survival’ (v3.7) on the clustered patients. Kaplan-Meier survival curves from diagnosis were calculated using census date 01.07.2022 with right censoring, along with Cox proportional hazard models adjusting for age at diagnosis and sex. Survival analysis underwent bootstrap repeat sampling using the R package ‘boot’ (v1.3) for censored data, using 1000 independent replicates. Clinical differences measured at point of diagnosis were compared between clusters.

### Detection of cell surface BMPR-II using spectral flow cytometry in leucocytes

Preliminary testing of two antibodies to the BMPR-II ECD (Thermo IgG1 3F6F8 and R&D Systems IgG2B 73805) were performed by western blot against a purified peptide of the BMPR-II ECD (*42*) and flow cytometry of PBMCs (Fig. SM2A-B). The R&D Systems IgG2B was selected for further use and was subsequently conjugated to the PE fluorophore using PE/R-Phycoerythrin Conjugation Kit - Lightning-Link® (Abcam) at a concentration of 1mg/ml. Quantification of cell surface BMPR-II staining was gated against a fluorescence-minus-one staining control and geometric mean fluorescence intensity (gMFI) was recorded in the positive population. Isotype IgG2B (mouse, R&D Systems) was performed alongside BMPR-II staining and showed reduced binding compared to BMPRII IgG2B (Fig. SM2C), however the isotype was not used as gating control due to the documented high levels of non-specific binding and poor use as gating controls (*43*). Quantibrite PE beads (BD Bioscience) were used to standardize quantitation between runs by interpolating gMFI of positive population to a standard curve and reporting cell surface BMPR-II as PE molecules per cell. Standard curves of Quantibrite beads showed good comparability between runs (Fig. SM2D). Longitudinal samples for PAH patients and healthy controls were stained for BMPR-II and a broad immunophenotyping panel for white blood cells and analysed in tandem using a Cytek Aurora spectral cytometer. Details of antibodies used, staining protocol and immunophenotyping gating strategy can be found in the supplemental methods. For analysis of flow cytometry data, results were adjusted for batch effect using linear modelling fluorescence MFI values using the R software. Batch differences were due to changes in BMPR-II antibody lot and conjugation to PE fluorophore. Residuals after batch correction for both PAH patients and healthy controls underwent consensus clustering using partition around medoids, Kmeans and hierarchical clustering, using the R package ‘Consensus Cluster Plus’ using 1000 repetitions, and parameters set at maxK=6, pItem = 0.8 and Euclidian distances. Kmeans clustering K=3 was found to be optimal. Levels of cell surface BMPR-II on each cell type were compared between clusters and Kruskal-Wallis used to assess statistical significance.

### Statistical analysis

Statistical analysis was performed using GraphPad Prism version 10 and R Studio. The normality of all variables was tested using the Shapiro-Wilk test. Normally distributed variables are presented as mean ± standard deviation and compared using Students t-test or ANOVA. Non-normally distributed variables are presented as median ± interquartile range (IQR) and compared using the Wilcoxon test for two paired groups or Kruskal-Wallis test with Dunn’s multiple comparison testing. A p<0.05 was considered statistically significant. All statistical tests were two sided. If applicable, multiple testing correction was applied using FDR with the Benjami-Hochberg procedure. Spearman’s rank correlation co-efficients were determined using the Rpackage ‘Hmisc’ and plotted using the ‘ggplot’ function.

## Code availability

Code is available at GitHub rjd58/StratosPHere1

## Data availability

The transcriptomic and clinical data used in this study have been deposited in the EGA (the European Genome-phenome Archive) database under accession code EGAS00001005532 and are accessible as previously referenced (*41*)

## List of Supplementary Materials

**Materials and Methods**

StratosPHere 1 Biomarker Study: Collection and Processing of blood samples.

Supporting methodology for MIQE guidelines of quantitative PCR.

Additional supplementary methods:

External biomarker qPCR validation.

Immunophenotyping and BMPR-II quantification in leukocyte populations using spectral flow cytometry.

Culture of THP1 cells.

Table SM1 Primers for genes of interest and reference genes. Table SM2. cDNA and primer concentrations used in qPCR.

Table SM3: Antibodies used for spectral flow cytometry immunophenotyping for the analysis of cell surface BMPR-II in major leucocyte populations.

Fig.SM1. Development of house-keeping genes for qPCR in whole blood samples in PAH. Fig.SM2. Development of detection of cell surface BMPR-II using flow cytometry.

Fig.SM3. Gating strategy for immunophenotyping and detection of cell surface BMPR-II in leukocyte populations.

**Results**

Table S1. Temporal stability of candidate biomarker GOIs tested in the StratosPHere 1 biomarker study.

Table S2. Demographics and clinical characteristics of patients and healthy volunteers used in the RNAseq analysis of the UK National Cohort Study of Idiopathic and Heritable Pulmonary Arterial Hypertension.

Table S3. Spearman’s rank correlation coefficients and p-values matrices for biomarker panel genes

Table S4. Biomarker gene expression between unsupervised machine learning clusters of RNAseq data

Table S5. Clinical characteristics of patients stratified unsupervised machine learning clustering of biomarker panel using RNAseq values

Table S6. Demographics and clinical characteristics of patients and healthy volunteers used in the qPCR validation cohort.

Table S7. Cell surface BMPR-II levels following clustering analysis.

Table S8. Plasma proteomic data of TGFβ associated proteins measured in PAH.

Fig. S1. Supporting results for development of the transcriptomic BMPR-II biomarker panel.

Fig. S2. Longitudinal RNAseq analysis of Biomarker Panel genes in PAH patients recruited measured as part of A) the UK PAH cohort study and B) TRANSFORM-UK trial.

Fig. S3. Supporting results for unsupervised machine learning clustering of the BMPR-II biomarker panel using RNAseq data from the UK PAH cohort study.

Fig. S4. Validation cohort for the BMPR-II biomarker panel.

Fig. S5. Supporting results for BMPR-II cell surface staining in leukocyte populations using samples from StratosPHere 1.

Fig. S6. THP1 monocytic cell line treated with BMP ligands.

## Acknowledgments

We thank the patients who have generously donated time to help with our research. This funding was supported by MRC grant (MR/T025646/1). E.M.D.D.D. was funded by Gates Scholarship. The UK National Cohort of Idiopathic and Heritable PAH is supported by the National Institute of Health Research BioResource, the BHF (SP/12/12/29836), and the UK Medical Research Council (MR/K020919/1). Author contributions: R.J.J. and M.T. conceived of and designed the study. R.J.J. performed experiments and analyzed data. E.M.D.D.D, A.Y.K.N., B.D., N.D., S.V. and P.D.U. provided technical support. G.P. and J.N. recruited patients to the study. E.M.D.D.D., C.R., A.L. and S.G. provided UK National PAH cohort data. J.W. and A.H. provided validation cohort. R.J.J. and M.T. drafted and revised the manuscript.

## References

1. Y. C. Lai, K. C. Potoka, H. C. Champion, A. L. Mora, M. T. Gladwin, Pulmonary arterial hypertension: The clinical syndrome. Circulation Research 115, 115–130 (2014).

2. P. D. Upton, N. W. Morrell, TGF-β and BMPR-II pharmacology-implications for pulmonary vascular diseases. Current Opinion in Pharmacology 9, 274–280 (2009).

3. L. Yung, P. Yang, S. Joshi, Z. M. Augur, S. S. J. Kim, G. A. Bocobo, T. Dinter, L. Troncone, P. Chen, M. E. Mcneil, M. Southwood, S. P. De Frias, J. Knopf, I. O. Rosas, D. Sako, R. S. Pearsall, J. D. Quisel, G. Li, R. Kumar, P. B. Yu, ACTRIIA-Fc rebalances activin / GDF versus BMP signaling in pulmonary hypertension. SCIENCE TRANSLATIONAL MEDICINE 12 (2020).

4. L. Southgate, R. D. Machado, S. Gräf, N. W. Morrell, Molecular genetic framework underlying pulmonary arterial hypertension. Nature Reviews Cardiology 17, 85–95 (2020).

5. R. C. Trembath, J. R. Thomson, R. D. Machado, N. V. Morgan, C. Atkinson, I. Winship, G. Simonneau, N. Galie, J. E. Loyd, M. Humbert, W. C. Nichols, J. Berg, A. Manes, J. McGaughran, M. Pauciulo, L. Wheeler, N. W. Morrell, Clinical and Molecular Genetic Features of Pulmonary Hypertension in Patients with Hereditary Hemorrhagic Telangiectasia. New England Journal of Medicine 345, 325–334 (2001).

6. R. E. Harrison, R. Berger, S. G. Haworth, R. Tulloh, C. J. Mache, N. W. Morrell, M. A. Aldred, R. C. Trembath, Transforming growth factor-β receptor mutations and pulmonary arterial hypertension in childhood. Circulation 111, 435–441 (2005).

7. M. T. Nasim, T. Ogo, M. Ahmed, R. Randall, H. M. Chowdhury, K. M. Snape, T. Y. Bradshaw, L. Southgate, G. J. Lee, I. Jackson, G. M. Lord, J. S. R. Gibbs, M. R. Wilkins, K. Ohta-Ogo, K. Nakamura, B. Girerd, F. Coulet, F. Soubrier, M. Humbert, N. W. Morrell, R. C. Trembath, R. D. Machado, Molecular genetic characterization of SMAD signaling molecules in pulmonary arterial hypertension. Human Mutation 32, 1385–1389 (2011).

8. S. Gräf, M. Haimel, M. Bleda, C. Hadinnapola, L. Southgate, W. Li, J. Hodgson, B. Liu, R. M. Salmon, M. Southwood, R. D. Machado, J. M. Martin, C. M. Treacy, K. Yates, L. C. Daugherty, O. Shamardina, D. Whitehorn, S. Holden, M. Aldred, H. J. Bogaard, C. Church, G. Coghlan, R. Condliffe, P. A. Corris, C. Danesino, M. Eyries, H. Gall, S. Ghio, H. A. Ghofrani, J. S. R. Gibbs, B. Girerd, A. C. Houweling, L. Howard, M. Humbert, D. G. Kiely, G. Kovacs, R. V. MacKenzie Ross, S. Moledina, D. Montani, M. Newnham, A. Olschewski, H. Olschewski, A. J. Peacock, J. Pepke-Zaba, I. Prokopenko, C. J. Rhodes, L. Scelsi, W. Seeger, F. Soubrier, D. F. Stein, J. Suntharalingam, E. M. Swietlik, M. R. Toshner, D. A. Van Heel, A. Vonk Noordegraaf, Q. Waisfisz, J. Wharton, S. J. Wort, W. H. Ouwehand, N. Soranzo, A. Lawrie, P. D. Upton, M. R. Wilkins, R. C. Trembath, N. W. Morrell, Identification of rare sequence variation underlying heritable pulmonary arterial hypertension. Nature Communications 9 (2018).

9. T. D. Mueller, J. Nickel, Promiscuity and specificity in BMP receptor activation. FEBS Letters 586, 1846–1859 (2012).

10. C. Guignabert, L. Savale, A. Boucly, R. Thuillet, L. Tu, M. Ottaviani, C. J. Rhodes, P. De Groote, G. Prévot, E. Bergot, A. Bourdin, L. S. Howard, E. Fadel, A. Beurnier, A. Roche, M. Jevnikar, X. Jaïs, D. Montani, M. R. Wilkins, O. Sitbon, M. Humbert, Serum and Pulmonary Expression Profiles of the Activin Signaling System in Pulmonary Arterial Hypertension. Circulation 147, 1809–1822 (2023).

11. M. M. Hoeper, D. B. Badesch, H. A. Ghofrani, J. S. R. Gibbs, M. Gomberg-Maitland, V. V. McLaughlin, I. R. Preston, R. Souza, A. B. Waxman, E. Grünig, G. Kopeć, G. Meyer, K. M. Olsson, S. Rosenkranz, Y. Xu, B. Miller, M. Fowler, J. Butler, J. Koglin, J. de Oliveira Pena, M. Humbert, Phase 3 Trial of Sotatercept for Treatment of Pulmonary Arterial Hypertension. New England Journal of Medicine 388, 1478–1490 (2023).

12. L. Long, X. Yang, M. Southwood, J. Lu, S. J. Marciniak, B. J. Dunmore, N. W. Morrell, Chloroquine prevents progression of experimental pulmonary hypertension via inhibition of autophagy and lysosomal bone morphogenetic protein type II receptor degradation. Circulation Research 112, 1159–1170 (2013).

13. B. J. Dunmore, K. M. Drake, P. D. Upton, M. R. Toshner, M. A. Aldred, N. W. Morrell, The lysosomal inhibitor, chloroquine, increases cell surface BMPR-II levels and restores BMP9 signalling in endothelial cells harbouring BMPR-II mutations. Human Molecular Genetics 22, 3667–3679 (2013).

14. B. J. Dunmore, X. D. Yang, A. Crosby, S. Moore, L. Long, C. Huang, M. Southwood, E. D. Austin, A. Rana, P. D. Upton, N. W. Morrell, 4Pba Restores Signaling of a Cysteine-Substituted Mutant Bmpr2 Receptor Found in Patients With Pulmonary Arterial Hypertension. American Journal of Respiratory Cell and Molecular Biology 63, 160–171 (2020).

15. L. Long, M. L. Ormiston, X. Yang, M. Southwood, S. Gräf, R. D. Machado, M. Mueller, B. Kinzel, L. M. Yung, J. M. Wilkinson, S. D. Moore, K. M. Drake, M. A. Aldred, P. B. Yu, P. D. Upton, N. W. Morrell, Selective enhancement of endothelial BMPR-II with BMP9 reverses pulmonary arterial hypertension. Nature Medicine 21, 777–785 (2015).

16. S. A. Bustin, V. Benes, J. A. Garson, J. Hellemans, J. Huggett, M. Kubista, R. Mueller, T. Nolan, M. W. Pfaffl, G. L. Shipley, J. Vandesompele, C. T. Wittwer, The MIQE guidelines: Minimum information for publication of quantitative real-time PCR experiments. Clinical Chemistry 55, 611–622 (2009).

17. N.. D. R.. M. A.. S. J.. D. M.. J. R.. W. M.. T. M.. V. S. Deliu, StratosPHere 2: A response-adaptive randomised placebo-controlled Phase II trial to evaluate hydroxychloroquine and phenylbutyrate in pulmonary arterial hypertension caused by mutations in BMPR2. *Pre-Print* (2023).

18. C. J. Rhodes, P. Otero-Núñez, J. Wharton, E. M. Swietlik, S. Kariotis, L. Harbaum, M. J. Dunning, J. M. Elinoff, N. Errington, A. A. R. Thompson, J. Iremonger, J. G. Coghlan, P. A. Corris, L. S. Howard, D. G. Kiely, C. Church, J. Pepke-Zaba, M. Toshner, S. J. Wort, A. A. Desai, M. Humbert, W. C. Nichols, L. Southgate, D. A. Trégouët, R. C. Trembath, I. Prokopenko, S. Gräf, N. W. Morrell, D. Wang, A. Lawrie, M. R. Wilkins, Whole-blood RNA profiles associated with pulmonary arterial hypertension and clinical outcome. American Journal of Respiratory and Critical Care Medicine 202, 586–594 (2020).

19. M. Toshner, C. Church, L. Harbaum, C. Rhodes, S. S. Villar Moreschi, J. Liley, R. Jones, A. Arora, K. Batai, A. A. Desai, J. G. Coghlan, S. J. R. Gibbs, D. Gor, S. Gräf, L. Harlow, J. Hernandez-Sanchez, L. S. Howard, M. Humbert, J. Karnes, D. G. Kiely, R. Kittles, E. Knightbridge, B. Lam, K. A. Lutz, W. C. Nichols, M. W. Pauciulo, J. Pepke-Zaba, J. Suntharalingam, F. Soubrier, R. C. Trembath, T. H. L. Schwantes-An, S. John Wort, M. R. Wilkins, S. Gaine, N. W. Morrell, P. A. Corris, Mendelian randomisation and experimental medicine approaches to interleukin-6 as a drug target in pulmonary arterial hypertension. European Respiratory Journal 59 (2022), doi:10.1183/13993003.02463-2020.

20. N. Rudarakanchana, J. A. Flanagan, H. Chen, P. D. Upton, R. Machado, D. Patel, R. C. Trembath, N. W. Morrell, Functional analysis of bone morphogenetic protein type II receptor mutations underlying primary pulmonary hypertension. Human Molecular Genetics 11, 1517– 1525 (2002).

21. G. Pousada, V. Lupo, S. Cástro-Sánchez, M. Álvarez-Satta, A. Sánchez-Monteagudo, A. Baloira, C. Espinós, Di. Valverde, Molecular and functional characterization of the BMPR2 gene in Pulmonary Arterial Hypertension. Scientific Reports 7, 1–19 (2017).

22. C. Atkinson, S. Stewart, P. D. Upton, R. Machado, J. R. Thomson, R. C. Trembath, N. W. Morrell, Primary pulmonary hypertension is associated with reduced pulmonary vascular expression of type II bone morphogenetic protein receptor. Circulation 105, 1672–1678 (2002).

23. C. J. Rhodes, J. Wharton, E. M. Swietlik, L. Harbaum, B. Girerd, J. G. Coghlan, J. Lordan, C. Church, J. Pepke-Zaba, M. Toshner, S. J. Wort, D. G. Kiely, R. Condliffe, A. Lawrie, S. Gräf, D. Montani, A. Boucly, O. Sitbon, M. Humbert, L. S. Howard, N. W. Morrell, M. R. Wilkins, Using the Plasma Proteome for Risk Stratifying Patients with Pulmonary Arterial Hypertension. American Journal of Respiratory and Critical Care Medicine 205, 1102–1111 (2022).

24. T. A. Samad, A. Rebbapragada, E. Bell, Y. Zhang, Y. Sidis, S. J. Jeong, J. A. Campagna, S. Perusini, D. A. Fabrizio, A. L. Schneyer, H. Y. Lin, A. H. Brivanlou, L. Attisano, C. J. Woolf, DRAGON, a bone morphogenetic protein co-receptor. Journal of Biological Chemistry 280, 14122–14129 (2005).

25. J. L. Babitt, Y. Zhang, T. A. Samad, Y. Xia, J. Tang, J. A. Campagna, A. L. Schneyer, C. J. Woolf, H. Y. Lin, Repulsive guidance molecule (RGMa), a DRAGON homologue, is a bone morphogenetic protein co-receptor. Journal of Biological Chemistry 280, 29820–29827 (2005).

26. J. L. Babitt, F. W. Huang, D. M. Wrighting, Y. Xia, Y. Sidis, T. A. Samad, J. A. Campagna, R. T. Chung, A. L. Schneyer, C. J. Woolf, N. C. Andrews, H. Y. Lin, Bone morphogenetic protein signaling by hemojuvelin regulates hepcidin expression. Nature Genetics 38, 531–539 (2006).

27. M. Moser, O. Binder, Y. Wu, J. Aitsebaomo, R. Ren, C. Bode, V. L. Bautch, F. L. Conlon, C. Patterson, BMPER, a Novel Endothelial Cell Precursor-Derived Protein, Antagonizes Bone Morphogenetic Protein Signaling and Endothelial Cell Differentiation. Molecular and Cellular Biology 23, 5664–5679 (2003).

28. S. Sudo, O. Avsian-Kretchmer, L. S. Wang, A. J. W. Hsueh, Protein related to DAN and cerberus is a bone morphogenetic protein antagonist that participates in ovarian paracrine regulation. Journal of Biological Chemistry 279, 23134–23141 (2004).

29. S. Aykul, E. Martinez-Hackert, Transforming growth factor-β family ligands can function as antagonists by competing for type II receptor binding. Journal of Biological Chemistry 291, 10792–10804 (2016).

30. J. C. Bendell, M. S. Gordon, H. I. Hurwitz, S. F. Jones, D. S. Mendelson, G. C. Blobe, N. Agarwal, C. H. Condon, D. Wilson, A. E. Pearsall, Y. Yang, T. McClure, K. M. Attie, M. L. Sherman, S. Sharma, Safety, pharmacokinetics, pharmacodynamics, and antitumor activity of dalantercept, an activin receptor-like kinase-1 ligand trap, in patients with advanced cancer. Clinical Cancer Research 20, 480–489 (2014).

31. P. D. Upton, R. J. Davies, R. C. Trembath, N. W. Morrell, Bone morphogenetic protein (BMP)and activin type II receptors balance BMP9 signals mediated by activin receptor-like kinase-1 in human pulmonary artery endothelial cells. Journal of Biological Chemistry 284, 15794–15804 (2009).

32. L. Long, M. Ormiston, X. Yang, M. Southwood, S. Graf, R. Machado, M. Mueller, B. Kinzel, L. Yung, J. Wilkinson, S. Moore, K. Drake, M. Adlred, P. Yu, P. Upton, N. Morrell, Selective enhancement of endothelial BMPR-II with BMP9 reverses pulmonary arterial hypertension. Nature Medicine 21, 777–785 (2015).

33. L. A. Hurst, B. J. Dunmore, L. Long, A. Crosby, R. Al-Lamki, J. Deighton, M. Southwood, X. Yang, M. Z. Nikolic, B. Herrera, G. J. Inman, J. R. Bradley, A. A. Rana, P. D. Upton, N. W. Morrell, TNFα drives pulmonary arterial hypertension by suppressing the BMP type-II receptor and altering NOTCH signalling. Nature Communications 8 (2017), doi:10.1038/ncomms14079.

34. A. Babicheva, J. X. J. Yuan, Endothelial Notch1 in Pulmonary Hypertension: Friend or Foe? Circulation Research 124, 176–179 (2019).

35. K. Miyagawa, M. Shi, P. I. Chen, J. K. Hennigs, Z. Zhao, M. Wang, C. G. Li, T. Saito, S. Taylor, S. Sa, A. Cao, L. Wang, M. P. Snyder, M. Rabinovitch, Smooth Muscle Contact Drives Endothelial Regeneration by BMPR2-Notch1-Mediated Metabolic and Epigenetic Changes. Circulation Research 124, 211–224 (2019).

36. Y. Xing, S. Zhao, Q. Wei, S. Gong, X. Zhao, F. Zhou, R. AI-Lamki, D. Ortmann, M. Du, R. Pedersen, G. Shang, S. Si, N. W. Morrell, J. Yang, A novel piperidine identified by stem cell-based screening attenuates pulmonary arterial hypertension by regulating BMP2 and PTGS2 levels. European Respiratory Journal 51 (2018), doi:10.1183/13993003.02229-2017.

37. T. Katagiri, T. Watabe, Bone morphogenetic proteins. Cold Spring Harbor Perspectives in Biology 8, 1–27 (2016).

38. T. Helbing, R. Rothweiler, E. Ketterer, L. Goetz, J. Heinke, S. Grundmann, D. Duerschmied, C. Patterson, C. Bode, M. Moser, BMP activity controlled by BMPER regulates the proinflammatory phenotype of endothelium. Blood 118, 5040–5049 (2011).

39. E. Cahill, C. M. Costello, S. C. Rowan, S. Harkin, K. Howell, M. O. Leonard, M. Southwood, E. P. Cummins, S. F. Fitzpatrick, C. T. Taylor, N. W. Morrell, F. Martin, P. McLoughlin, Gremlin plays a key role in the pathogenesis of pulmonary hypertension. Circulation 125, 920–930 (2012).

40. M. Toshner, C. Church, L. Harbaum, C. Rhodes, S. S. V. Moreschi, J. Liley, R. Jones, A. Arora, K. Batai, A. A. Desai, J. G. Coghlan, J. S. R. Gibbs, D. Gor, S. Gräf, L. Harlow, J. Hernandez-sanchez, L. S. Howard, M. Humbert, J. Karnes, D. G. Kiely, R. Kittles, E. Knightbridge, B. Lam, K. A. Lutz, W. C. Nichols, M. W. Pauciulo, J. Pepke-zaba, J. Suntharalingam, F. Soubrier, R. C. Trembath, S. J. Wort, M. Wilkins, S. Gaine, N. W. Morrell, A. Paul, Early View Mendelian randomisation and experimental medicine approaches to IL-6 as a drug target in PAH. European Respiratory Journal (2021).

41. S. Kariotis, E. Jammeh, E. M. Swietlik, J. A. Pickworth, C. J. Rhodes, P. Otero, J. Wharton, J. Iremonger, M. J. Dunning, D. Pandya, T. S. Mascarenhas, N. Errington, A. A. R. Thompson, C. E. Romanoski, F. Rischard, J. G. N. Garcia, J. X.-J. Yuan, T.-H. S. An, A. A. Desai, G. Coghlan, J. Lordan, P. A. Corris, L. S. Howard, R. Condliffe, D. G. Kiely, C. Church, J. Pepke-Zaba, M. Toshner, S. Wort, S. Gräf, N. W. Morrell, M. R. Wilkins, A. Lawrie, D. Wang, M. Bleda, C. Hadinnapola, M. Haimel, K. Auckland, T. Tilly, J. M. Martin, K. Yates, C. M. Treacy, M. Day, A. Greenhalgh, D. Shipley, A. J. Peacock, V. Irvine, F. Kennedy, S. Moledina, L. MacDonald, E. Tamvaki, A. Barnes, V. Cookson, L. Chentouf, S. Ali, S. Othman, L. Ranganathan, J. S. R. Gibbs, R. DaCosta, J. Pinguel, N. Dormand, A. Parker, D. Stokes, D. Ghedia, Y. Tan, T. Ngcozana, I. Wanjiku, G. Polwarth, R. V. Mackenzie Ross, J. Suntharalingam, M. Grover, A. Kirby, A. Grove, K. White, A. Seatter, A. Creaser-Myers, S. Walker, S. Roney, C. A. Elliot, A. Charalampopoulos, I. Sabroe, A. Hameed, I. Armstrong, N. Hamilton, A. M. K. Rothman, A. J. Swift, J. M. Wild, F. Soubrier, M. Eyries, M. Humbert, D. Montani, B. Girerd, L. Scelsi, S. Ghio, H. Gall, A. Ghofrani, H. J. Bogaard, A. V. Noordegraaf, A. C. Houweling, A. H. in’t Veld, G. Schotte, Biological heterogeneity in idiopathic pulmonary arterial hypertension identified through unsupervised transcriptomic profiling of whole blood. Nature Communications 12, 1–14 (2021).

42. H. Jiang, R. M. Salmon, P. D. Upton, Z. Wei, A. Lawera, A. P. Davenport, N. W. Morrell, W. Li, The prodomain-bound form of bone morphogenetic protein 10 is biologically active on endothelial cells. Journal of Biological Chemistry 291, 2954–2966 (2016).

43. M. N. Andersen, S. N. H. Al-Karradi, T. W. Kragstrup, M. Hokland, Elimination of erroneous results in flow cytometry caused by antibody binding to Fc receptors on human monocytes and macrophages. Cytometry Part A 89, 1001–1009 (2016).

44. G. Monaco, B. Lee, W. Xu, S. Mustafah, Y. Y. Hwang, C. Carré, N. Burdin, L. Visan, M. Ceccarelli, M. Poidinger, A. Zippelius, J. Pedro de Magalhães, A. Larbi, RNA-Seq Signatures Normalized by mRNA Abundance Allow Absolute Deconvolution of Human Immune Cell Types. Cell Reports 26, 1627–1640.e7 (2019).

