## Supplementary File for "BMPR-II biomarkers for testing therapeutic efficacy in pulmonary arterial hypertension –the StratosPHere 1 study"

### Contents

**Supplementary Methods**

Supplementary methods: Supplementary methods for external validation cohort

Supplementary methods: Immunophenotyping and BMPR-II quantification in leukocyte populations using spectral flow cytometry

Supplementary methods: Culture of THP1 cells

Table 1. SM1: qPCR primers.

Table SM2: cDNA and primer concentrations used in qPCR.

Table SM3: Antibodies used for spectral flow cytometry immunophenotyping for the analysis of cell surface BMPR-II in major leucocyte populations.

Fig. SM1. Development of house-keeping genes for qPCR in whole blood samples in PAH.

Fig. SM2. Development of detection of cell surface BMPR-II using flow cytometry.

Fig. SM3. Gating strategy for immunophenotyping and detection of cell surface BMPR-II in leukocyte populations.

**Supplementary Results**

Table S1. Temporal stability of candidate biomarker GOIs tested in the StratosPHere 1 biomarker study.

Table S2. Demographics and clinical characteristics of patients and healthy volunteers used in the RNAseq analysis of the UK National Cohort Study of Idiopathic and Heritable Pulmonary Arterial Hypertension.

Table S3. Spearman's rank correlation coefficients and p-values matrices for biomarker panel genes.

Table S4. Biomarker gene expression between unsupervised machine learning clusters of RNAseq data.

Table S5. Clinical characteristics of patients stratified unsupervised machine learning clustering of biomarker panel using RNAseq values.

Table S6. Demographics and clinical characteristics of patients and healthy volunteers used in the qPCR validation cohort.

Table S7. Cell surface BMPR-II levels following clustering analysis.

Table S8. Plasma proteomic data of TGF$\beta$ associated proteins measured in PAH.

Fig. S1. Supporting results for development of the transcriptomic BMPR-II biomarker panel.

Fig. S2. Longitudinal RNAseq analysis of Biomarker Panel genes in PAH patients recruited measured as part of the UK PAH cohorts study and TRANSFORM-UK trial.

Fig. S3. Supporting results for unsupervised machine learning clustering of the BMPR-II biomarker panel using RNAseq data from the UK PAH cohort study.

Fig. S4. Validation cohort of our BMPR2 biomarker panel.

Fig. S5. Supporting results for BMPR-II cell surface staining in leukocyte populations using samples from StratosPHere 1.

Fig. S6. THP1 monocytic cell line treated with BMP ligands.

Supplementary methods: StratosPHere I Biomarker Study: Collection and Processing of blood samples.

Supplementary methods: Supplementary methods for measurement of transcriptomic biomarkers in whole blood

### Supplementary Methods

#### StratosPHere I Biomarker Study: Collection and Processing of blood samples.

**Tempus RNA tubes**

Whole blood was collected into Tempus$^{TM}$ RNA tubes (ThermoFisher Scientific) according to manufacturer's instructions and stored at -80°C until processing. Isolation of RNA and cDNA synthesis was performed in batches with all donor timepoints prepared together to reduce potential batch variation. All steps were performed according to manufacturer's instructions unless otherwise stated. RNA was isolated from whole blood using the TEMPUS RNA spin isolation kit (ThermoFisher Scientific). A total of two Tempus$^{TM}$ tubes per donor timepoint were pooled. Isolated RNA was subsequently treated with DNase to remove contaminating genomic DNA (DNA-free kit, ThermoFisher Scientific). Finally, due to the nature of the Tempus$^{TM}$ blood RNA extraction kit, the resultant RNA is of high volume, low concentration and so a subsequent concentration step using RNeasy MinElute columns (Qiagen) was performed. RNA samples were analysed for concentration and A260/280 using a Nanodrop Spectrophotometer (ThermoFisher Scientific).

**Isolation of Peripheral Blood Mononuclear cells**

Whole blood was collected into ACD-A tubes (Greiner) and subsequently diluted 1:1 in D-PBS (without Mg2+ or Ca2+, Sigma) and then fractioned using Leucosep tubes (Greiner) containing Ficoll Paque PLUS reagent (Cytiva) according to manufacturer's instructions. Isolated white blood cells were removed and subjected to red blood cell lysis (Abcam) and washed three times in D-PBS (without Mg2+ or Ca2+) containing 2mM EDTA (ThermoFisher Scientific) and centrifuging at 300xg for 5 minutes between each wash. Washed and pelleted cells were stored in a medium of 90% foetal bovine serum (Sigma) containing 10% DMSO (Sigma) in liquid nitrogen.

**Plasma and Serum**

Plasma samples were isolated from EDTA tubes (Greiner) following centrifugation at 1800xg for 10 minutes. Serum was isolated from CAT Serum Sep clot activator tubes following 30 minute incubation in an upright position and following centrifugation at 1800xg for 10 minutes. Samples were stored at -80°c.

#### Supporting methodology for MIQE guidelines of quantitative PCR

To be in line with MIQE guidelines for the development of whole blood RNA biomarkers the following additional information is given.

**Nucleic Acid Extraction**: Purity of RNA was measured post extraction using the Tempus$^{TM}$ RNA isolation spin kit and post concentration using RNeasy MinElute columns (Qiagen). RNA Purity and quantification were measured using NanoDrop with an A260/280 ratio between 1.9-2.1 as acceptable. A final yield of >140ng/μl was expected.
**Reverse transcription**: This was performed using 1400ng of RNA in a 20μl reaction.
**qPCR target information**: Primer sequences along with accession number (including additional variants) and amplicon length are given in table SM1. *In silico* specificity was confirmed using BLAST (NCBI).
**qPCR protocol**: Whole blood cDNA samples were serially diluted to identify optimal sample reaction concentration. Sample cDNA was added at a volume of 2μl in a final reaction volume of 10μl. Where primers were designed in house, concentration is given. See Table SM2. Standard curves for the purpose of qPCR and validation and relative gene expression were included on plates. Standard curved were generated using either cDNA derived from pulmonary arterial endothelial cells (PAECs) treated with 1ng/ml BMP9 treatment for 4 hours following an overnight serum starvation with media containing 0.1% foetal bovine serum or from cDNA from

Tempus whole blood, depending on where expression was the highest to gain the best possible information. Calibration curves for primer sets were performed using log fold dilutions (10ng – 0.01ng) of sample to assess PCR efficiency using the formula PCR efficiency = $10^{(-1/slope)} - 1$, when the logarithm of the initial template concentration is plotted on the x axis and the Cq is plotted on the y axis and slope of the curve. A PCR efficiency of 100% ±10% and an $R^2 > 0.98$ was deemed acceptable. Melt curves were analysed for single peak products and no template / reverse transcription controls for non-specific amplification where values were accepted if undetermined of >10Ct above samples.

**qPCR analysis:** Ct values for genes of interest were read off a cycle threshold set in the linear phase of the log(ΔRN) vs cycle number, well before the reaction plateaued with a baseline correction set at cycles 6-12. For multiple plates assessing the same gene, the cycle threshold was kept constant. Average Ct values for samples were calculated, and outlier technical replicates >0.5Ct were excluded from final analysis (excluding repeatability calculations). Samples were normalised by two methods:

1. Standard curve method: Ct values of logarithmic (base10) standard curves were plotted using GraphPad Prism and simple linear regression performed. Sample Ct values were then interpolated into the standard curve to generate an interpolated value which was converted to ng with log normalisation and multiplied by the cDNA dilution factor. Gene of interest data values were normalised to the mean of 4 reference genes (*B2M, GAPDH, YWHAZ* and *HPRT*) (gene of interest divided by mean reference gene). Fold change from baseline to week 16 was calculated for each donor.
2. ΔΔCt method: Ct values for samples were normalised to the mean Ct value of 4 reference genes (*B2M, GAPDH, YWHAZ and HPRT-1*) and relative gene expression fold change between time points determined using the ΔΔCt method.

**Development of house-keeping genes for whole blood assays:** Reference genes for the normalisation of qPCR data were also assessed in whole blood samples. 10 candidate house-keeping genes were assessed for expression in whole blood RNA with a pre-defined cut-off of ≤Ct 25, of which three were excluded (*PBGD, RPL27* and *TBP*, Fig.SM1A). Seven were tested in StratosPHere 1 PAH patient samples (N=20) and 2 excluded due to high variation amongst PAH donors (*EEF1a1* and *EEF2*, Fig.SM1B). *B2M, GAPDH, HPRT, PGK1* and *YWHAZ* were assessed and for assay repeatability (Fig.SM1C) and reproducibility (Fig.SM1D). Four reference genes were selected for qPCR normalisation purposes (*B2M, GAPDH, HPRT-1* and *YWHAZ*) and 1 gene (*PGK1*) kept in reserve should treatment affect any of the chosen genes within a clinical trial. The 5 housekeeping genes were also assessed for temporal stability, and comparison of Ct values over the time frame showed little variation (Fig.SM1E).

### <u>Additional supplementary methods</u>

#### External biomarker qPCR validation

10ng cDNA was amplified by qPCR using SYBR green reagents in a 20µl reaction using matched primers with the exception of *NOTCH1* (F: TGCCCTGGGCTTCTCTGG, R: TGGCACGATTTCCCTGACC) and *GAPDH* (F: GCC AAA TAT GAT GAC ATC AAG AAG G, R: GGT GTC GCT GTT GAA GTC AGA G) using the amplification protocol: 95°c for 10 minutes; followed by 40 cycles of 95°c for 15 seconds, 60°c for 1 minute; followed by 95°c for 15 seconds, 60°c for 1 minute, 95°c for 15 seconds melt curve on a QuantStudio3 (ThermoFisher Scientific). ΔCT calculation for biomarker gene expression were determined after normalisation to 4 housekeeping genes (*B2M, GAPDH, HPRT-1 and YWHAZ*) as described in main methods.

#### Immunophenotyping and BMPR-II quantification in leukocyte populations using spectral flow cytometry

Thawed PBMC aliquots were washed in D-PBS, centrifuged at 600xg for 5 minutes and resuspended at 1 million cells per 100µl. Cells were then blocked with 5µl human TruStain FcX Fc receptor block (Biolegend) per 1million cells at room temperature for 10 minutes. A test volume refers to 1 million cells in100µl volume. Per test, cells were incubated for 30 minutes at 4°C with 1µl (R&D Systems) BMPR-II antibody at 1mg/ml that had been conjugated to PE using the PE / R-Phycoerythrin Conjugation Kit - Lightning-Link® (Abcam) kit. After 30 minutes, the test samples were subsequently incubated with an immunophenotyping stain complex as described in Table SM3 for a further 30 minutes at 4°C with the inclusion of propidium iodide for dead cell exclusion. Stained samples were washed 2x in cell staining buffer (Biolegend) and centrifuged at 600xg for 5 minutes between washes, before resuspending in 200µl. Single staining controls were used to train the Cytek Aurora for the purpose of spectral unmixing for each of the fluorescent tags and was performed on a regular basis. Immunophenotyping gating was performed as shown in Fig.SM3) for the identification of major white blood cell populations. A minimum of 100,000 live events were collected. FMO samples were run for the purpose of gating BMPR-II. Longitudinal samples for donor samples were stained and analysed in duplicate. Subsequent analysis was performed using FlowJo version 10.

**Culture of THP1 cells**

THP1 cells were routinely cultured in RPMI (Gibco) containing 10% foetal bovine serum, 10mM HEPES and 10mM sodium pyruvate (Sigma). Ligand stimulations were preceded by overnight serum depletion in media supplemented with 0.1% foetal bovine serum.

**Table SM1** Primers for genes of interest and reference genes are listed alphabetically, Qiagen QuantiTect primers are a proprietary sequence. In house primers were designed using NCBI Primer Blast with a required amplicon length of 70-200bp and span an exon-exon junction.

| Gene | Full name | Accession number | Amplicon length (bp) | Primer sequence |
|---|---|---|---|---|
| ARL4C | ADP ribosylation factor like GTPase 4C | NM_001282431.2 | 109 | F: GAAGAAGCGGACTGGTGACT<br>R: AGCTCTGGCGTTCAGACAAA |
| ACTB | B actin | NM_001101.5 | 70 | F: ACAGAGCCTCGCCTTTGCC<br>R: GATATCATCATCCATGGTGAGCTGG |
| APLN | Apelin | NM_017413.5 | 145 | F-CCAGAGGGTCAAGGAATGGGC<br>R-ATAACCGCCGGGGGTGGGCA |
| B2M | Beta-2-microglobulin | NM_004048.4 | 85 | F: CTCGCGCTACTCTCTCTTTCT<br>R: CATTCTCTGCTGGATGACGTG |
| CHSY3 | Chondroitin sulphate synthase 3 | NM_175856.5 | 72 | F: AGTAGGAAGATGCGTTCGCC<br>R: TGGAACAGTTGTTGCATCTC |
| DLX2 | Distal-less homeobox 2 | NM_004405.4 | 72 | F: GTCCCTTACTCCGCCAAGAG<br>R: TCCATAGGGAGCGTAGGAGG |
| EEF1a1 | Eukaryotic translation elongation factor 1 alpha 1 | NM_001402.6 | 194 | F: TATTGGATTGCCACACGGCT<br>R: AAAGCGACCCAAAGGTGGAT |
| EEF2 | Eukaryotic translation elongation factor 2 | NM_001961.4 | 140 | F: AAGTCCACGCTGACAGACTC<br>R: AAGAGGGAGATGGCAGTTGAC |
| EML6 | EMAP like 6 | NM_001039753.4 | 114 | F: AAGCTCCAAGAGGCAAACGG<br>R: TATGTGCTGGCCAGATTCCC |

| | | | | |
|---|---|---|---|---|
| FGFR3 | Fibroblast growth factor receptor 3 | NM_000142.5<br>NM_001354809.2 | 111 | F: GCACAACGTCACCTTTGAGG<br>R: AGCTCCTCCTCGGCTGG |
| GAPDH | Glyceraldehyde-3-phosphate dehydrogenase | NM_002046.7<br>NM_001289745.3<br>NM_001289746.2 | 181 | Qiagen QuantiTect: QT00079247 |
| HEY1 | Hes related family bHLH transcription factor with YRPW motif 1 | NM_001040708.1 | 126 | Qiagen QuantiTect: QT00035644 |
| HEY 2 | Hes related family bHLH transcription factor with YRPW motif 2 | NM_012259.3 | 131 | F: GCCATACAGATGCCGACAGA<br>R: CAGTTACCGAGCTGCCTTGA |
| HPRT1 | Hypoxanthine phosphoribosyltransferase 1 | NM_000194.3 | 140 | F:GCTATAAATTCTTTGCTGACCTGCTG<br>R: AATTACTTTTATGTCCCCTGTTGACTGG |
| ID1 | Inhibitor of DNA binding 1 | NM_002165 | 127 | Qiagen QuantiTect: QT00230650 |
| ID2 | Inhibitor of DNA binding 2 | NM_002166.5 | 112 | F: GACCCGATGAGCCTGCTATAC<br>R: GGTGCTGCAGGATTTCCATCT |
| ID3 | Inhibitor of DNA binding 3 | NM_002167 | 131 | F: CCCACCTTCCCATCCAGACAG<br>R: CCGAGTCAGTGGCAAAAGCTC |
| ID4 | Inhibitor of DNA binding 4 | NM_001546 | 149 | Qiagen QuantiTect: QT00234920 |
| KIT | KIT proto-oncogene, receptor tyrosine kinase | NM_000222.3 | 183 | F: TACTTGGAGCCTGCACCATT<br>R: TATCGCTGCAGGAAGACTCC |
| Notch 1 | Notch receptor 1 | NM_017617 | 108 | Qiagen QuantiTect: QT00231056 |
| Notch 2 | Notch receptor 2 | NM_024408 | 141 | F:CCAGGCTATCAGGGTGTCAAC<br>R: TTCACAGAGTAGGCCCCGAGT |
| PBGD | Hydroxymethylbilane synthase | NM_000190.4 | 130 | F: ATTACCCCGGGAGACTGAAC<br>R: GGCTGTTGCTTGGACTTCTC |
| PGK1 | Phosphoglycerate kinase 1 | NM_000291.4 | 158 | F: TGGGAACAAGGTTAAAGCCGA<br>R: AAAACCCACCAGCCTTCTGT |
| PTGS2 | Prostaglandin-endoperoxide synthase 2 | NM_000963.4 | 116 | F: GCTGTTCCCACCCATGTCAA<br>R: AAATTCCGGTGTTGAGCAGT |
| RAI2 | Retinoic acid induced 2 | NM_001172743.2<br>NM_001172739.2 | 146 | F: CCTTTGGGGGAAGAGGATCAC<br>R: ACTTGGCCTTCCAAGGAGTA |
| RPL27 | Ribosomal protein L27 | NM_000988.5 | 116 | F: GGGTGGTTGCTGCCGAA<br>R: GGTGCCATCATCAATGTTCTTCAC |
| Smad 1 | SMAD family member 1 | NM_005900 | 148 | F: TAGAAAGCCCTGTACTTCCTC<br>R: GGTTGCTGGAAAGAATCTGG |
| Smad 5 | SMAD family member 5 | NM_001001420<br>NM_001001419<br>NM_005903 | 139 | F: GAGAGTCCAGTCTTACCTCC<br>R: GGAAAGAATCTGGAAACGTG |
| Smad 6 | SMAD family member 6 | NM_005585 | 135 | Qiagen QuantiTect: QT00025242 |
| Smad 7 | SMAD family member 7 | NM_005904 | 123 | Qiagen QuantiTect: QT00076391 |
| SPRY1 | Sprouty RTK signaling antagonist 1 | NM_005841 | 85 | Qiagen QuantiTect: QT00219898 |

| | | | | |
|---|---|---|---|---|
| TBP | TATA-box binding protein | NM_003194.5 | 85 | F: GTGACCCAGCATCACTGTTTC<br>R: GAGCATCTCCAGCACACTCT |
| YWHAZ | Tyrosine 3-monooxygenase/tryptophan 5-monooxygenase activation protein zeta | NM_145690.3 | 141 | F: ACTTGACATTGTGGACATCGGA<br>R: GTGGGACAGCATGGATGACA |

**Table SM2**. cDNA and primer concentrations used in qPCR. Primer concentrations were determined for all in house designed primers and give the best signal:noise ratio. Standard curves were run with all genes of interest, either using endothelial cell sample (PAEC) or Tempus whole blood.

| **Gene** | **cDNA final quantity** | **Primer Concentration (forward and reverse)** | **Standard Curve used** |
|---|---|---|---|
| *ARL4C* | 7ng | Tempus whole blood | Tempus whole blood |
| *ID2* | 7ng | 800nM | PAEC + 1ng/ml BMP9 |
| *ID3* | 14ng | 800nM | PAEC + 1ng/ml BMP9 |
| *NOTCH1* | 7ng | N/A - Qiagen | PAEC + 1ng/ml BMP9 |
| *NOTCH2* | 7ng | 800nM | PAEC + 1ng/ml BMP9 |
| *PTGS2* | 7ng | 800nM | PAEC + 1ng/ml BMP9 |
| *SMAD 1* | 14ng | 400nM | PAEC + 1ng/ml BMP9 |
| *SMAD 5* | 14ng | 400nM | PAEC + 1ng/ml BMP9 |
| *SMAD 7* | 7ng | N/A - Qiagen | PAEC + 1ng/ml BMP9 |
| *SPRY1* | 14ng | N/A - Qiagen | PAEC + 1ng/ml BMP9 |
| *B2M* | 7ng | 800nM | Tempus whole blood |
| *EEF1a1* | 7ng | 800nM | PAEC + 1ng/ml BMP9 |
| *EEF2* | 7ng | 800nM | PAEC + 1ng/ml BMP9 |
| *GAPDH* | 7ng | N/A - Qiagen | PAEC + 1ng/ml BMP9 |
| *HPRT* | 14ng | 400nM | PAEC + 1ng/ml BMP9 |
| *PGK1* | 7ng | 400nM | PAEC + 1ng/ml BMP9 |
| *YWHAZ* | 7ng | 800nM | PAEC + 1ng/ml BMP9 |

**Table SM3:** Antibodies used for spectral flow cytometry immunophenotyping for the analysis of cell surface BMPR-II in major leucocyte populations.

| Fluorophore | Marker | Clone | Volume Per Test (100ul/1M) | Product ID | Supplier |
|---|---|---|---|---|---|
| BV421 | CD14 | M5E2 | 2.5 | 301829 | Biolegend |
| BV510 | CD45 | 2D1 | 2.5 | 368526 | Biolegend |
| BV650 | CD3 | UCHT1 | 2.5 | 300468 | Biolegend |
| BV711 | CD123 | 6H6 | 5 | 306030 | Biolegend |
| BV750 | CD56 | 5.1H11 | 2.5 | 362556 | Biolegend |
| AF488 | CD11c | Bu15 | 5 | 337236 | Biolegend |
| AF647 | HLADR | L243 | 2.5 | 307622 | Biolegend |
| SPARK YG 593 | CD19 | HIB19 | 2.5 | 302279 | Biolegend |
| APC Fire 810 | CD16 | 3G8 | 2.5 | | Biolegend |
| PI | PI | | 0.2 | | |

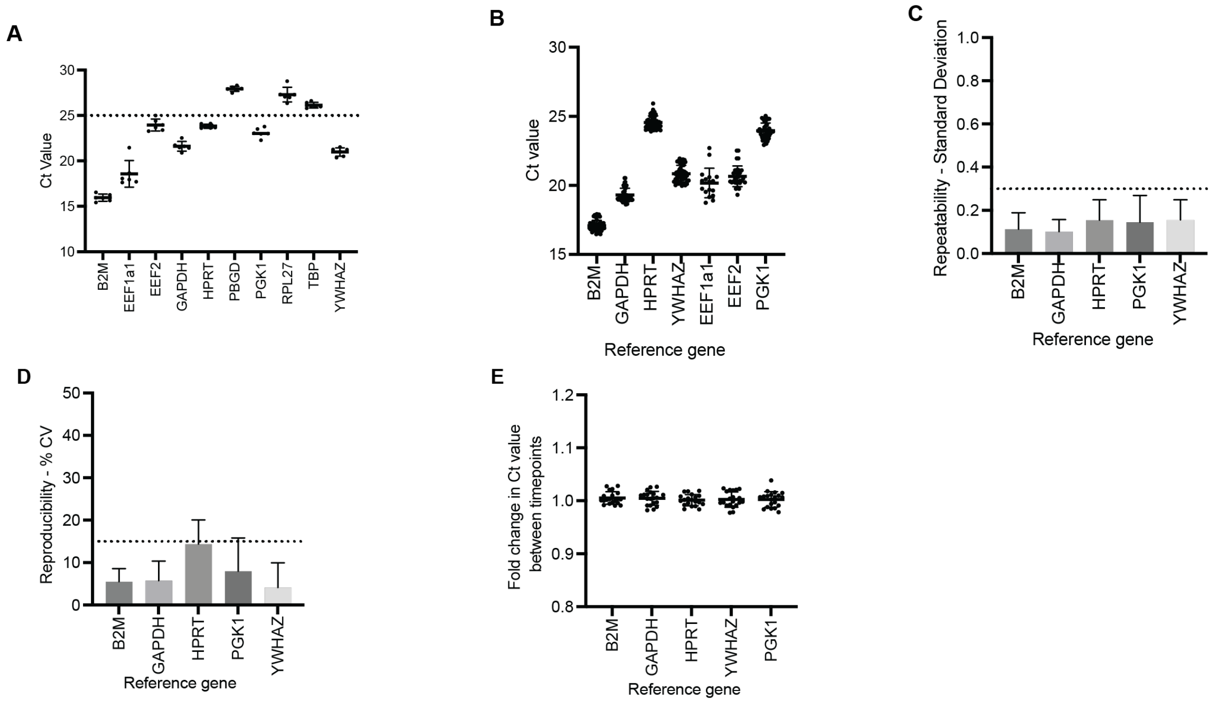


**Fig.SM1**. **Development of house-keeping genes for qPCR in whole blood samples in PAH. (A)** Preliminary screening for the detection of endogenous housekeeping genes required for qPCR normalisation from whole blood TEMPUS RNA (N=6, 6ng cDNA. Ct readout is given as an indicator of gene expression. A cut-off of Ct <25 was used for candidate biomarker genes (indicated by dotted lines). Data shown as mean ± standard deviation. **(B)** Ct values house-keeping genes in StratosPHere 1 samples (minimum, N=17) showing spread of values between donors. Data shown as mean ± SD. vC) **(C)** Repeatability of house-keeping gene Ct values measured across quadruplet replicates and reported as mean standard deviation on a per gene basis. Data shown as mean standard deviation ± SD. A pre-defined cut-off of maximum 0.3SD was applied (shown as dotted line). **(D)** Reproducibility of house-keeping gene Ct values as measured in a minimum of 8 sample repeats to assess accuracy and reported as % co-efficient of variation on a per gene basis. Data shown as mean % CV ± SD. A pre-defined cut-off of maximum 15% CV was applied (shown as dotted line). **(E)** Temporal stability of Ct values of house-keeping genes in PAH patients (N=20) between baseline and week 16 and reported as fold change on a per gene basis. Data shown as mean ± SD with dotted lines at ±0.3 fold change for assessment purposes.

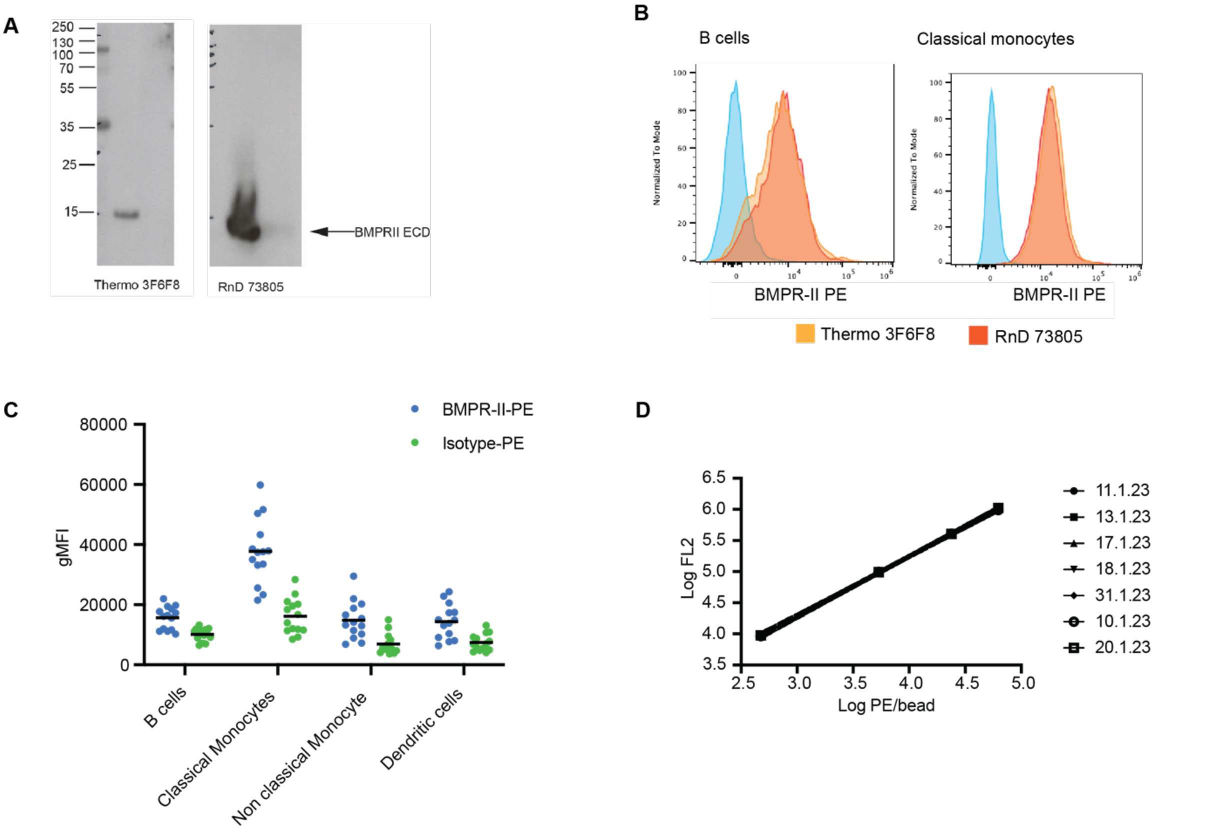


**Fig.SM2**. **Development of detection of cell surface BMPR-II using flow cytometry. (A)** Binding of antibodies raised against the ECD epitope of BMPRII to a peptide of BMPRII ECD. A recombinant peptide of the BMPRII ECD was probed by western blotting with commercial antibodies raised against the ECD; Thermo 3F6F8 clone probed against 5ug ECD and R&D Systems 73805 clone probed against 1ug ECD. The expected molecular weight of the ECD is 13.8kD. **(B)** Cell surface staining histograms of BMPR-II in representative B cell and classical monocyte populations using the Thermo 3F6F8 clone (orange) and R&D Systems 73805 clone (red), unstained shown as blue. **(C)** Comparison of cell surface staining in multiple leucocyte populations using either R&D Systems BMPR-II antibody conjugated to PE or IgG2B matched isotype (R&D Systems) conjugated to PE (N=12) data shown as gMFI with mean. **(D)** Standard curves generated using Quantibrite PE bead (BD Bioscience) standard curves show good consistency of PE detection between runs on the Cytek Aurora. **(E)** Immunophenotyping gating strategy for the detection of major leucocyte populations used for the quantification of cell surface BMPR-II.

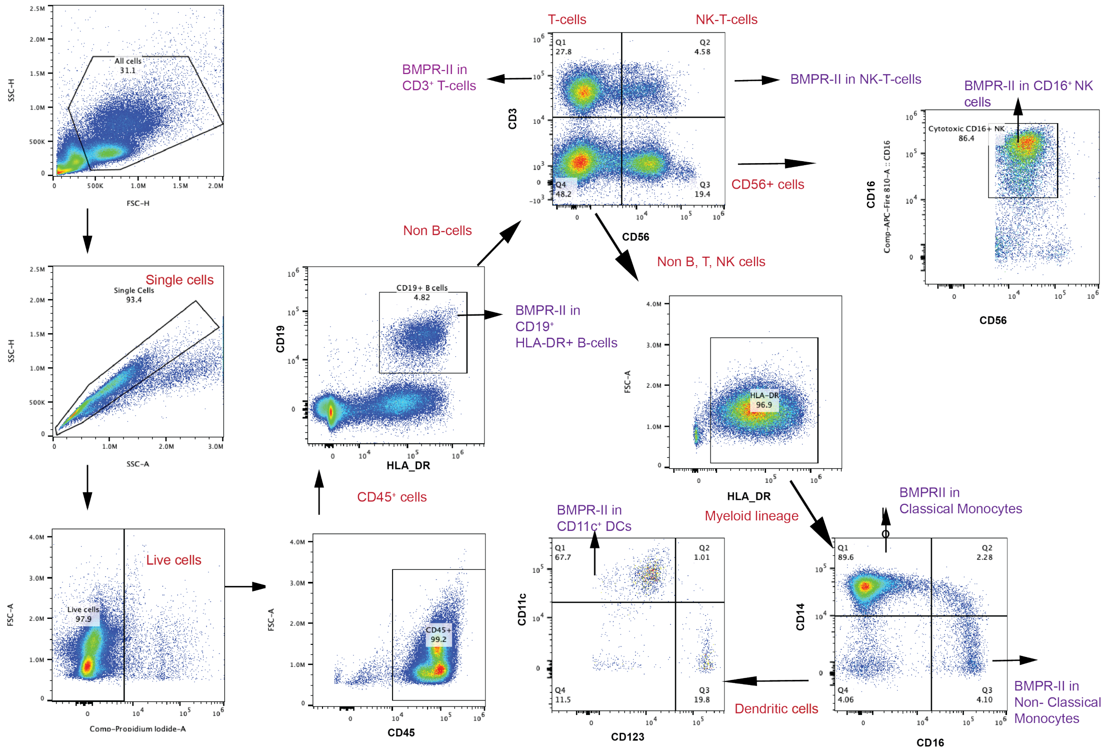


BMPR-II is detected in the PE channel

**Fig.SM3. Gating strategy for immunophenotyping and detection of cell surface BMPR-II in leukocyte populations.** Stained PBMCs were first separated by FSCvsSSC, then identification of single cells and then live/dead exclusion using propidium iodide. Broad classes of white blood cell were then detected as shown, with BMPR-II being detected in the PE channel.

### Supplemental Results

**Table S1. Temporal stability of candidate biomarker GOIs tested in the StratosPHere 1 biomarker study.** PAH patients (N=20) and controls (N=15) were sampled at baseline and 16 weeks (± 4 weeks). RNA isolated from whole blood. qPCR was used to test temporal stability of GOIs using two methods: Standard Curve and ΔΔCT. Table shows mean temporal fold change and standard deviation (SD) of fold change (FC) in each population and the mean % co-efficient of variation (%CV) between baseline and 16 weeks for each population.

| Gene | Population | Standard Curve Method | | | ΔΔCT Method | | |
|---|---|---|---|---|---|---|---|
| | | Mean FC | SD of FC | Mean %CV | Mean FC | SD of FC | Mean %CV |
| ***ARL4C*** | Control | 1.138 | 0.423 | 24.600 | 1.107 | 0.368 | 23.529 |
| | PAH | 1.026 | 0.194 | 9.884 | 1.029 | 0.216 | 11.335 |
| ***ID2*** | Control | 1.049 | 0.168 | 10.077 | 1.031 | 0.184 | 10.646 |
| | PAH | 0.984 | 0.159 | 8.788 | 0.983 | 0.170 | 9.492 |
| ***ID3*** | Control | 1.097 | 0.374 | 21.816 | 1.071 | 0.336 | 19.567 |
| | PAH | 1.128 | 0.310 | 15.828 | 1.129 | 0.312 | 15.398 |
| ***NOTCH1*** | Control | 1.026 | 0.292 | 13.286 | 1.011 | 0.275 | 16.338 |
| | PAH | 1.045 | 0.276 | 14.650 | 1.055 | 0.327 | 16.651 |
| ***NOTCH2*** | Control | 1.109 | 0.541 | 20.179 | 1.073 | 0.428 | 19.547 |
| | PAH | 1.041 | 0.222 | 11.870 | 1.046 | 0.243 | 12.905 |
| ***PTGS2*** | Control | 1.355 | 0.701 | 23.974 | 1.322 | 0.606 | 24.141 |
| | PAH | 0.995 | 0.425 | 21.098 | 1.015 | 0.489 | 22.535 |
| ***SMAD1*** | Control | 1.152 | 0.288 | 11.770 | 1.128 | 0.224 | 11.120 |
| | PAH | 1.062 | 0.161 | 9.392 | 1.065 | 0.176 | 9.416 |
| ***SMAD5*** | Control | 1.137 | 0.377 | 13.047 | 1.093 | 0.240 | 10.804 |
| | PAH | 0.999 | 0.210 | 8.779 | 0.997 | 0.220 | 10.510 |

**Table S2. Demographics and clinical characteristics of patients and healthy volunteers used in the RNAseq analysis of the UK National Cohort Study of Idiopathic and Heritable Pulmonary Arterial Hypertension.** Abbreviations: PAWP – pulmonary arterial wedge pressure; mPAP – mean pulmonary arterial pressure; mRAP – mean right atrial pressure; PVR – pulmonary vascular resistance; 6MWD – six minute walk distance; NT-proBNP – N-terminal pro b-type natriuretic peptide.

| | | **PAH patients** | **Healthy volunteers** |
|---|---|---|---|
| Number (IPAH / BMPR2-PAH) | | 356 (285/71) | 67 |
| Sex (Count) | Female | 252 | 43 |
| | Male | 104 | 24 |
| Age (at diagnosis, mean, range) | | 47 (11.7-79.3) | 43 (19.01-87.7) |
| Years since diagnosis sampled (Mean ± SD) | | 5.32 (4.92) | |
| PAWP (mmHg) (Mean, SD) | | 9.6 (3.8) | |
| | missing | 48 | |
| mPAP (mmHg) (Mean, SD) | | 54.8 (13.8) | |
| | missing | 14 | |
| mRAP (mmHg) (Mean, SD) | | 9.5 (5.3) | |
| | missing | 30 | |
| Cardiac output (l/min) (Mean, SD) | | 4.1 (1.4) | |
| | missing | 26 | |
| PVR (dynes/sec/cm$^{-5}$) (Mean, SD) | | 999.7 (480.9) | |
| | missing | 41 | |
| Cardiac Index (l/min/m$^2$) (Mean, SD) | | 2.2 (0.8) | |
| | missing | 222 | |
| 6MWD (meters) (Mean, SD) | | 302(140) | |
| | missing | 143 | |
| NT-pro-BNP (pg/ml) (Mean, SD) | | 1,510.7 (1,785.3) | |
| | missing | 252 | |
| Functional Class WHO I (Count, %) | WHO I | 2 (0.6) | |
| | WHO II | 62 (18.2) | |
| | WHO III | 236(69.4) | |
| | WHO IV | 40 (11.8) | |
| | missing | 15 | |

**Table S3**. **Spearman's rank correlation coefficients and p-values matrices for biomarker panel genes (A)** controls (N=67) **(B)** PAH (N=356). Black = Spearman's rank correlation co-efficient, blue = p-value.

A)

| | ARL4C | ID2 | ID3 | NOTCH1 | NOTCH2 | PTGS2 | SMAD1 | SMAD5 |
|---|---|---|---|---|---|---|---|---|
| **ARL4C** | 1.00 | 0.79 | 0.62 | -0.32 | -0.26 | -0.55 | 0.00 | 0.60 |
| **ID2** | <0.0001 | 1.00 | 0.34 | -0.17 | -0.10 | -0.37 | 0.03 | 0.62 |
| **ID3** | <0.0001 | 0.0055 | 1.00 | -0.34 | -0.31 | -0.49 | -0.2 | 0.22 |
| **NOTCH1** | 0.0092 | 0.175 | 0.0055 | 1.00 | 0.87 | 0.86 | 0.32 | 0.11 |
| **NOTCH2** | 0.034 | 0.4121 | 0.0097 | <0.0001 | 1.00 | 0.83 | 0.38 | 0.20 |
| **PTGS2** | <0.0001 | 0.0018 | <0.0001 | <0.0001 | <0.0001 | 1.00 | 0.23 | -0.02 |
| **SMAD1** | 0.999 | 0.6153 | 0.098 | 0.0087 | 0.0015 | 0.0628 | 1.00 | 0.18 |
| **SMAD5** | <0.0001 | <0.0001 | 0.0735 | 0.389 | 0.108 | 0.85 | 0.150 | 1.00 |

B)

| | ARL4C | ID2 | ID3 | NOTCH1 | NOTCH2 | PTGS2 | SMAD1 | SMAD5 |
|---|---|---|---|---|---|---|---|---|
| **ARL4C** | 1.00 | 0.79 | 0.56 | -0.20 | -0.06 | -0.33 | 0.09 | 0.63 |
| **ID2** | <0.0001 | 1.00 | 0.24 | -0.02 | 0.13 | -0.16 | 0.26 | 0.66 |
| **ID3** | <0.0001 | <0.0001 | 1.00 | -0.18 | -0.08 | -0.20 | -0.08 | 0.33 |
| **NOTCH1** | 0.00026 | 0.67 | <0.0001 | 1.00 | 0.80 | 0.71 | 0.28 | 0.12 |
| **NOTCH2** | 0.23 | 0.017 | 0.12 | <0.0001 | 1.00 | 0.64 | 0.50 | 0.30 |
| **PTGS2** | <0.0001 | 0.0022 | 0.0052 | <0.0001 | <0.0001 | 1.00 | 0.18 | -0.09 |
| **SMAD1** | 0.10 | <0.0001 | 0.16 | <0.0001 | <0.0001 | 0.00066 | 1.00 | 0.38 |
| **SMAD5** | <0.0001 | <0.0001 | <0.0001 | 0.020 | <0.0001 | 0.11 | <0.0001 | 1.00 |

**Table S4. Biomarker gene expression between unsupervised machine learning clusters of RNAseq data**. TPM RNAseq values for the BMPR-II biomarker panel genes had been clustered using data from the UK PAH cohort study using optimised Kmeans=3. Data shown as mean TPM ± SD. Overall ANOVA p value shown, with tukeys range test performed between clusters (**** p<0.0001, ** p<0.01, * p<0.05, ns = non-significant.

| Biomarker gene | Cluster 1 (N=146) | Cluster 2 (N=138) | Cluster 3 (N= 72) | Average (N=356) | p-value | 1v2 p-value | 1v3 p-value | 2v3 p-value |
|---|---|---|---|---|---|---|---|---|
| *ARL4C* | 41.5 (12) | 54 (15.5) | 24.2 (7.2) | 42.9 (16.8) | < 1e-04 | **** | **** | **** |
| *ID2* | 53.2 (15.9) | 59 (19.9) | 30.4 (9.3) | 50.8 (19.7) | < 1e-04 | * | **** | **** |
| *ID3* | 2.9 (1.2) | 3.9 (1.8) | 1.8 (0.8) | 3.1 (1.6) | < 1e-04 | **** | **** | **** |
| *NOTCH1* | 9.3 (4.1) | 5 (1.4) | 6.3 (2.5) | 7 (3.5) | < 1e-04 | **** | **** | * |
| *NOTCH2* | 78.2 (17) | 51 (9.8) | 52.6 (14.5) | 62.6 (19.2) | < 1e-04 | **** | **** | ns |
| *SMAD5* | 11.7 (2.6) | 11.4 (2.9) | 6.6 (1.8) | 10.5 (3.3) | < 1e-04 | ns | **** | **** |
| *SMAD1* | 3.5 (1) | 2.7 (0.9) | 2.4 (0.8) | 3 (1) | < 1e-04 | **** | **** | * |
| *PTGS2* | 11.4 (5.9) | 6.5 (2) | 9.1 (3.6) | 9 (4.8) | < 1e-04 | **** | *** | *** |

**Table S5. Clinical characteristics of patients stratified unsupervised machine learning clustering of biomarker panel using RNAseq values**. Data shown as count (%), mean±SD with Students t-test or median± IQR with Kruskal-Wallis performed depending on data normality following Shapiro-Wilk test. Abbreviations: PAWP – pulmonary arterial wedge pressure; mPAP – mean pulmonary arterial pressure; mRAP – mean right atrial pressure; PVR – pulmonary vascular resistance; 6MWD – six minute walk distance; NT-proBNP – N-terminal pro b-type natriuretic peptide.

| | | **Cluster 1** | **Cluster 2** | **Cluster 3** | **Total** | **p-value** | **FDR (q value)** |
|---|---|---|---|---|---|---|---|
| Number | | 146 | 138 | 72 | 356 | | |
| Sex (Count, %) | Female | 103 (70.5) | 93 (67.4) | 55 (76.4) | 251 (70.5) | | |
| | Male | 43 (29.5) | 45 (32.6) | 17 (23.6) | 105 (29.5) | 0.40 | 0.430 |
| Age at diagnosis (Mean, SD) | | 49.4 (14.5) | 44.6 (16.4) | 46.7 (14.5) | 47 (15.4) | 0.032 | 0.139 |
| BMPR2 mutation (Count, %) | No | 122 (83.6) | 111 (80.4) | 52 (72.2) | 285 (80.1) | 0.14 | 0.303 |
| | Yes | 24 (16.4) | 27 (19.6) | 20 (27.8) | 71 (19.9) | | |
| Years since diagnosis sampled (Mean ± SD) | | 5.53 (5.03) | 5.01 (4.53) | 5.73 (5.40) | 5.32 (4.92) | 0.85 | 0.85 |
| PAWP (mmHg) (Median, IQR) | | 10.0 (5.0) | 9.0 (5.0) | 10.0 (5.0) | 10.0 (5.0) | 0.22 | 0.317 |
| | missing | 15 | 21 | 13 | 49 | | |
| mPAP (mmHg) (Median, IQR) | | 53.5 (17.0) | 53.0 (18.0) | 55.0 (18.0) | 54.0 (17.0) | 0.37 | 0.43 |
| | missing | 4 | 6 | 5 | 15 | | |
| mRAP (mmHg) (Median, IQR) | | 8.0 (7.0) | 8.0 (6.0) | 10.0 (7.0) | 8.0 (6.0) | 0.06 | 0.156 |
| | missing | 12 | 12 | 7 | 31 | | |
| Cardiac output (l/min) (Median, IQR) | | 3.94 (1.73) | 3.97 (2.32) | 3.5 (1.81) | 3.8 (1.92) | 0.17 | 0.316 |
| | missing | 7 | 12 | 8 | 27 | | |
| PVR (dynes/sec/cm$^{-5}$) (Median, IQR) | | 899.3 (516.2) | 927.9 (721) | 1,083 (732.2) | 923.1 (626) | 0.20 | 0.318 |
| | missing | 14 | 18 | 10 | 42 | | |
| Cardiac Index (l/min/m$^2$) (Median, IQR) | | 2.1 (0.94) | 2.09 (1.26) | 1.67 (0.43) | 2.03 (0.96) | 0.053 | 0.156 |
| | missing | 95 | 77 | 51 | 223 | | |
| 6MWD (meters) (Median, IQR) | | 293 (210) | 355 (130) | 300 (219) | 320 (207.7) | 0.011 | 0.139 |
| | missing | 57 | 59 | 28 | 144 | | |
| NT-pro-BNP (pg/ml) (Median, IQR) | | 916.3 (2199.2) | 736.5 (1216.3) | 1541 (2981.0) | 867.5 (1997) | 0.3 | 0.39 |
| | missing | 102 | 104 | 47 | 253 | | |
| Functional Class WHO I (Count, %) | WHO I | 0 | 2(1.6) | 0 | 2 (0.6) | 0.026 | 0.139 |
| | WHO II | 20 (14.3) | 33 (25.6) | 9 (12.7) | 62 (18.2) | | |
| | WHO III | 101 (72.1) | 85 (65.9) | 50 (70.4) | 236(69.4) | | |
| | WHO IV | 19 (13.6) | 9 (7.0) | 12 (16.9) | 40 (11.8) | | |
| | missing | 5 | 9 | 1 | 15 | | |

**Table S6. Demographics and clinical characteristics of patients and healthy volunteers used in the qPCR validation cohort.** Abbreviations: mPAP – mean pulmonary arterial pressure; mRAP – mean right atrial pressure; PVR – pulmonary vascular resistance.

| | | PAH patients | Healthy volunteers |
|---|---|---|---|
| Number (IPAH/HPAH / CTD-PAH/other) | | 37(14/5/16/2) | 16 |
| Sex (Count) | Female | 17 | 10 |
| | Male | 10 | 6 |
| Age at sampling (Mean, range) | | 50.3 (24.8-78.9) | 35.4 (16.6-67.9) |
| mRAP (mmHg) (Mean, SD) | | 10.45 (5.7) | |
| mPAP (mmHg (Mean, SD) | | 59.6 (18.3) | |
| PVR (woods units) (Mean, SD) | | 12.5 (5.5) | |
| Cardiac Index (l/min/m$^2$) (Mean, SD) | | 1.86 (0.58) | |

**Table S7. Cell surface BMPR-II levels following clustering analysis (clusters A-C).** Cell surface BMPR-II levels following Kmeans K=3 cluster analysis of cell surface BMPR2 in B cells, monocytes and dendritic cells. Levels of cell surface BMPR-II and allocation of PAH patients (IPAH / *BMPR2*-PAH mutation carriers) and healthy controls. Group allocation shown as count (%) and cell surface BMPR2 shown as median PE molecules per cell ± IQR. Statistical analysis performed using Kruskal Wallis.

| | | Cluster A (N=15) | Cluster B (N=9) | Cluster C (N=21) | Total (N=45) | p value |
|---|---|---|---|---|---|---|
| **Group (count, %)** | **IPAH** | 9 (60%) | 4 (44.4%) | 7 (33.3%) | 20 (44.4%) | 0.053 |
| | **BMPR2-PAH** | 4 (26.7%) | 1 (11.1%) | 3 (14.3%) | 8 (17.8%) | |
| | **Control** | 2 (13.3%) | 4 (44.4%) | 11 (53.4%) | 17 (37.8%) | |
| **B cell (median, IQR)** | | 486.5 [376.7, 645.6] | 770.3 [419.6, 863.3] | 677.5 [504.5, 727.5] | 568.3 [401.6, 759.6] | 0.085 |
| **Classical monocyte (median, IQR)** | | 1,229.9 [1,112.1, 1,477.8] | 2,858.7 [2,486.3, 2,945.5] | 1,952.1 [1,776.9, 2,100.3] | 1,939.8 [1,495.6, 2,298.5] | <0.0001 |
| **Non classical monocyte (median, IQR)** | | 563.3 [391.0, 726.2] | 1,326.2 [791.9, 1,447.3] | 768.6 [649.1, 926.3] | 766.5 [584.9, 926.3] | 0.0003 |
| **Dendritic cell (median, IQR)** | | 579.1 [456.2, 631.3] | 1,210 [1,039.7, 1,671.5] | 882.1 [685.6, 1,058.1] | 819.8 [584.5, 1,058.1] | <0.0001 |

**Table S8. Plasma proteomic data of TGFβ associated proteins measured in PAH**. N=357 proteins measured as part of another study *(23)*. Estimate represents estimated difference in PAH versus healthy controls. Significance measured as FDR q value.

| Protein Full Name | Protein | Estimate | Standard Error | z value | Pr(>\|z\|) | FDR (q) |
|---|---|---|---|---|---|---|
| RGM domain family member B | RGMB | -0.743 | 0.100 | -7.459 | $8.89x10^{-14}$ | $6.36x10^{-08}$ |
| Repulsive guidance molecule A | RGMA | -0.708 | 0.109 | -6.525 | $6.78x10^{-11}$ | $1.14x10^{-08}$ |
| BMP-binding endothelial regulator protein | BMPER | 0.880 | 0.135 | 6.512 | $1.35x10^{-09}$ | $2.81x10^{-06}$ |
| Bone morphogenetic protein 10 | BMP10 | 0.597 | 0.099 | 6.062 | $3.58x10^{-09}$ | $1.21x10^{-05}$ |
| Follistatin-related protein 3 | FSTL3 | 0.735 | 0.125 | 5.902 | $7.33x10^{-08}$ | $3.09x10^{-06}$ |
| Transforming growth factor beta receptor type 3 | TGF-b R III | -0.531 | 0.099 | -5.383 | $8.44x10^{-08}$ | $3.31x10^{-05}$ |
| Hemojuvelin | RGMC | -0.708 | 0.132 | -5.357 | $8.72x10^{-08}$ | $7.91x10^{-05}$ |
| Inhibin beta B chain | Inhibin b B chain | 0.582 | 0.109 | 5.352 | $8.89x10^{-14}$ | 0.00014 |
| Gremlin-2 | GREM2 | 0.574 | 0.116 | 4.939 | $7.87x10^{-07}$ | 0.00044 |
| Growth/differentiation factor 11/8 | GDF-11/8 | -0.519 | 0.131 | -3.951 | $7.79x10^{-05}$ | 0.007 |
| Transforming growth factor beta-3 | TGF-b3 | 0.431 | 0.113 | 3.808 | 0.00014 | 0.010 |
| Inhibin beta A chain | Inhibin b A chain | 0.354 | 0.112 | 3.149 | 0.0016 | 0.028 |
| Bone morphogenetic protein 8B | BMP-8 | -0.296 | 0.107 | -2.758 | 0.006 | 0.072 |
| Inhibin beta C chain | INHBC | 0.299 | 0.116 | 2.588 | 0.010 | 0.045 |
| Noggin | Noggin | -0.238 | 0.093 | -2.575 | 0.010 | 0.136 |
| Bone morphogenetic protein receptor type-1B | BMP RIB | -0.229 | 0.093 | -2.458 | 0.014 | 0.167 |
| Bone morphogenetic protein 4 | BMP-4 | -0.288 | 0.130 | -2.213 | 0.027 | 0.589 |
| Bone morphogenetic protein 1 | BMP-1 | -0.227 | 0.107 | -2.121 | 0.034 | 0.211 |
| Left-right determination factor 2 | Lefty-A | -0.212 | 0.102 | -2.081 | 0.037 | 0.252 |
| Bone morphogenetic protein 7 | BMP-7 | 0.250 | 0.121 | 2.065 | 0.039 | 0.157 |
| Low-density lipoprotein receptor-related protein 1B | LRP1B | 0.252 | 0.126 | 1.995 | 0.046 | 0.187 |
| Serine/threonine-protein phosphatase PP1-gamma catalytic subunit | PP1G | 0.221 | 0.111 | 1.982 | 0.048 | 0.211 |
| Growth/differentiation factor 2 | GDF2 | -0.170 | 0.087 | -1.957 | 0.050 | 0.152 |
| Bone morphogenetic protein 4 | BMP-4 | 0.188 | 0.097 | 1.943 | 0.052 | 0.186 |
| Growth/differentiation factor 9 | GDF-9 | 0.187 | 0.096 | 1.942 | 0.052 | 0.279 |
| Prolow-density lipoprotein receptor-related protein 1 | LRP1 | -0.115 | 0.063 | -1.811 | 0.070 | 0.359 |
| Gremlin-1 | GREM1 | 0.249 | 0.142 | 1.751 | 0.080 | 0.213 |
| Mothers against decapentaplegic homolog 3 | SMAD3 | -0.129 | 0.074 | -1.743 | 0.081 | 0.199 |
| Activin A | Activin A | 0.182 | 0.109 | 1.668 | 0.095 | 0.209 |
| Activin receptor type-1B | Activin RIB | -0.237 | 0.161 | -1.475 | 0.140 | 0.819 |
| Mitogen-activated protein kinase 3 | ERK-1 | 0.151 | 0.109 | 1.382 | 0.167 | 0.169 |
| Follistatin-related protein 1 | FSTL1 | 0.105 | 0.089 | 1.177 | 0.239 | 0.846 |
| Bone morphogenetic protein 15 | BMP15 | 0.087 | 0.075 | 1.168 | 0.243 | 0.405 |
| Inhibin beta A chain:Inhibin beta B chain heterodimer | Activin AB | 0.101 | 0.088 | 1.156 | 0.248 | 0.281 |

| | | | | | | |
|---|---|---|---|---|---|---|
| Growth/differentiation factor 3 | GDF-3 | -0.127 | 0.120 | -1.065 | 0.287 | 0.780 |
| Mothers against decapentaplegic homolog 4 | SMAD4 | 0.084 | 0.081 | 1.034 | 0.301 | 0.686 |
| Follistatin | FST | 0.082 | 0.088 | 0.938 | 0.349 | 0.956 |
| Bone morphogenetic protein 6 | BMP-6 | 0.075 | 0.087 | 0.856 | 0.392 | 0.633 |
| Activin B | Activin B | -0.092 | 0.112 | -0.814 | 0.416 | 0.672 |
| Activin receptor type-2A | Activin RIIA | -0.058 | 0.074 | -0.777 | 0.437 | 0.576 |
| Bone morphogenetic protein 3 | BMP-3 | -0.059 | 0.079 | -0.740 | 0.460 | 0.335 |
| Mothers against decapentaplegic homolog 2 | SMAD2 | 0.058 | 0.081 | 0.715 | 0.475 | 0.601 |
| Left-right determination factor 2 | Lefty-A | 0.066 | 0.099 | 0.660 | 0.509 | 0.715 |
| Mitogen-activated protein kinase 9 | JNK2 | 0.050 | 0.076 | 0.659 | 0.510 | 0.855 |
| Inhibin beta C chain | INHBC | 0.060 | 0.094 | 0.633 | 0.527 | 0.359 |
| Deoxynucleoside triphosphate triphosphohydrolase SAMHD1 | SAMH1 | -0.053 | 0.088 | -0.609 | 0.543 | 0.706 |
| Follistatin-related protein 5 | FSTL5 | -0.041 | 0.069 | -0.593 | 0.553 | 0.642 |
| Transforming growth factor beta-2 | TGF-b2 | 0.067 | 0.120 | 0.552 | 0.581 | 0.733 |
| TGF-beta receptor type-2 | TGF-b R II | -0.050 | 0.097 | -0.516 | 0.606 | 0.916 |
| Transforming growth factor beta-1 | TGF-b1 | 0.064 | 0.129 | 0.492 | 0.623 | 0.714 |
| Growth/differentiation factor 5 | BMP-14 | 0.040 | 0.083 | 0.479 | 0.632 | 0.754 |
| Mothers against decapentaplegic homolog 1 | SMAD1 | -0.040 | 0.086 | -0.465 | 0.642 | 0.950 |
| Growth/differentiation factor 11 | GDF-11 | 0.045 | 0.114 | 0.394 | 0.694 | 0.658 |
| Bone morphogenetic protein receptor type-2 | BMP RII | 0.029 | 0.084 | 0.339 | 0.734 | 0.448 |
| Nodal homolog | NODAL | 0.031 | 0.104 | 0.302 | 0.763 | 0.639 |
| Inhibin beta A chain:Inhibin beta C chain heterodimer | Activin AC | 0.029 | 0.118 | 0.244 | 0.808 | 0.889 |
| Growth/differentiation factor 10 | BMP-3b | -0.021 | 0.087 | -0.243 | 0.808 | 0.530 |
| Activin receptor type-2B | Activin RIIB | -0.017 | 0.094 | -0.183 | 0.855 | 0.717 |
| Cerberus | CER1 | 0.013 | 0.109 | 0.118 | 0.906 | 0.773 |
| Bone morphogenetic protein 2 | BMP-2 | -0.012 | 0.135 | -0.088 | 0.930 | 0.721 |
| Growth/differentiation factor 7 | GDF7 | 0.004 | 0.078 | 0.045 | 0.964 | 0.720 |
| Growth/differentiation factor 10 | BMP-3b | -0.003 | 0.159 | -0.019 | 0.985 | 0.424 |
| Growth/differentiation factor 8 | Myostatin | 0.001 | 0.074 | 0.019 | 0.985 | 0.684 |

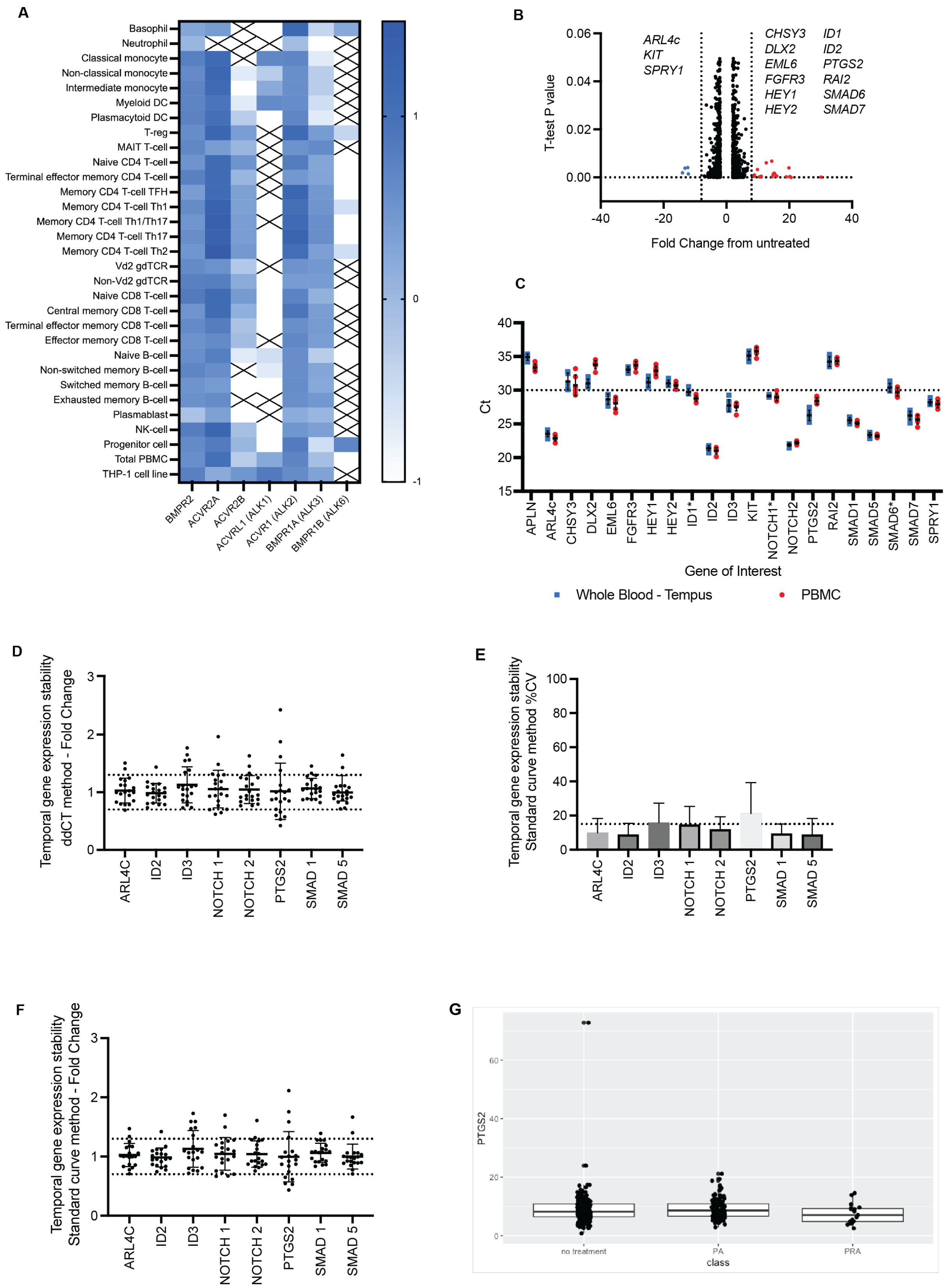


**Fig. S1. Supporting results for development of the transcriptomic BMPR-II biomarker panel. (A)** Transcript expression values for TGFβ family members; *BMPR2*, *ACRV2A, ACVR2B, ACVRL1, ACVR1, BMPR1A* and *BMPR1B*

from 29 blood immune cells as determined by Monaco *et al*. *(44)* and available on www.proteinatlas.org. Also shown for comparison is their expression in THP1 cells derived from the cell lines information on the website. RNAseq values were log scaled prior to presentation on the heatmap. Crosses represent no detectable expression. (**B)** Volcano plot showing genes proposed to be differentially regulated by BMPR-II in pulmonary arterial endothelial cells following stimulation with BMP9 ligand as reported in *(32)*. Dotted lines show a ± 7.5-fold change in expression. (**C)** Preliminary screening for the detection of candidate gene of interest biomarkers from whole blood TEMPUS RNA (N=6). cDNA was either assayed with 6ng or 40ng (denoted by a * on the gene name). Ct readout is given as an indicator of gene expression. A cut-off of Ct <30 was used for candidate biomarker genes (indicated by dotted lines). Comparison of readily detectable candidate biomarkers in both peripheral blood mononuclear cells (PBMCs, red) and Tempus whole blood (blue). Individual data points are shown for donor replicates, along with mean ± standard deviation. Note *ID4* was undetectable. (**D)** Temporal stability was assessed in PAH samples (N=20) between baseline and 16 weeks (± 4 weeks) using the $\Delta\Delta$CT method and shown as fold change mean fold change ± SD), dotted lines show ± 0.3-fold change. (**E)** Temporal stability was assessed in PAH samples (N=20) between baseline and 16 weeks (± 4 weeks) using the standard curve method and shown as % coefficient of variation dotted line at 15% CV. (**F)** Temporal stability was assessed in PAH samples (N=20) between baseline and 16 weeks (± 4 weeks) using the standard curve method and shown as fold change mean fold change ± SD), dotted lines show ± 0.3-fold change. (**G)** Effect of prostaglandin related medications upon *PTGS2* RNA expression as measured by RNAseq in PAH patients with; no relevant treatment = 224, PA = Epoprostenol, Iloprost, Treprostinil (N=115), PRA = Selexipag (N=17). Data shown as median ± IQR. Kruskal Wallis across groups p=0.1625.

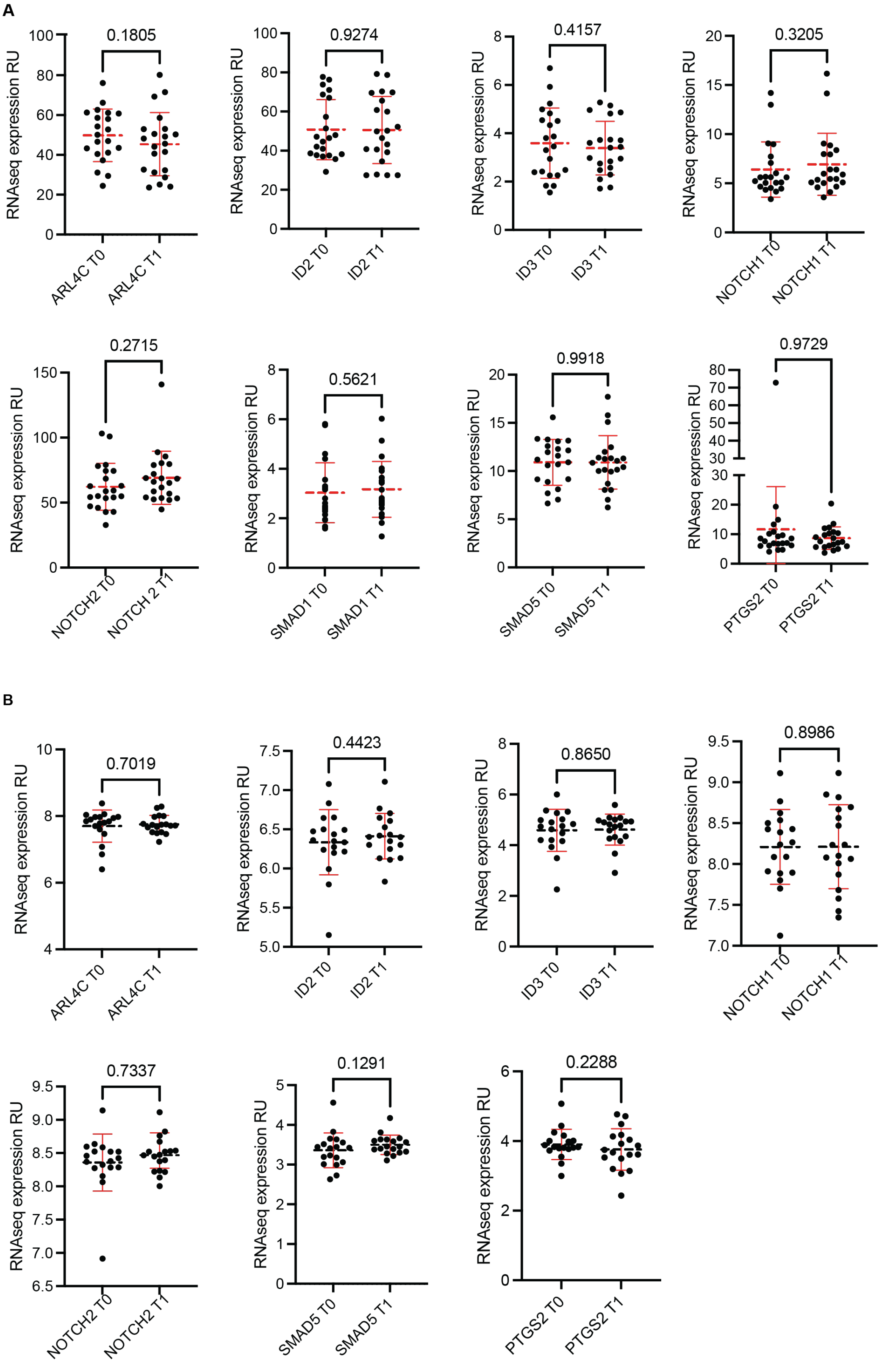


**Fig. S2. Longitudinal RNAseq analysis of Biomarker Panel genes in PAH patients recruited measured as part of A) the UK PAH cohort study and B) TRANSFORM-UK trial. (A)**PAH patients (N=21) compared longitudinally over a mean of 385 days (range: 50-1253 days), **(B)** PAH patients (N=18) compared longitudinally over 6 months. Data shown as individual data points mean ± SD in red bars. Statistical analysis between populations using paired T-test or Wilcoxon test, depending on data normality.

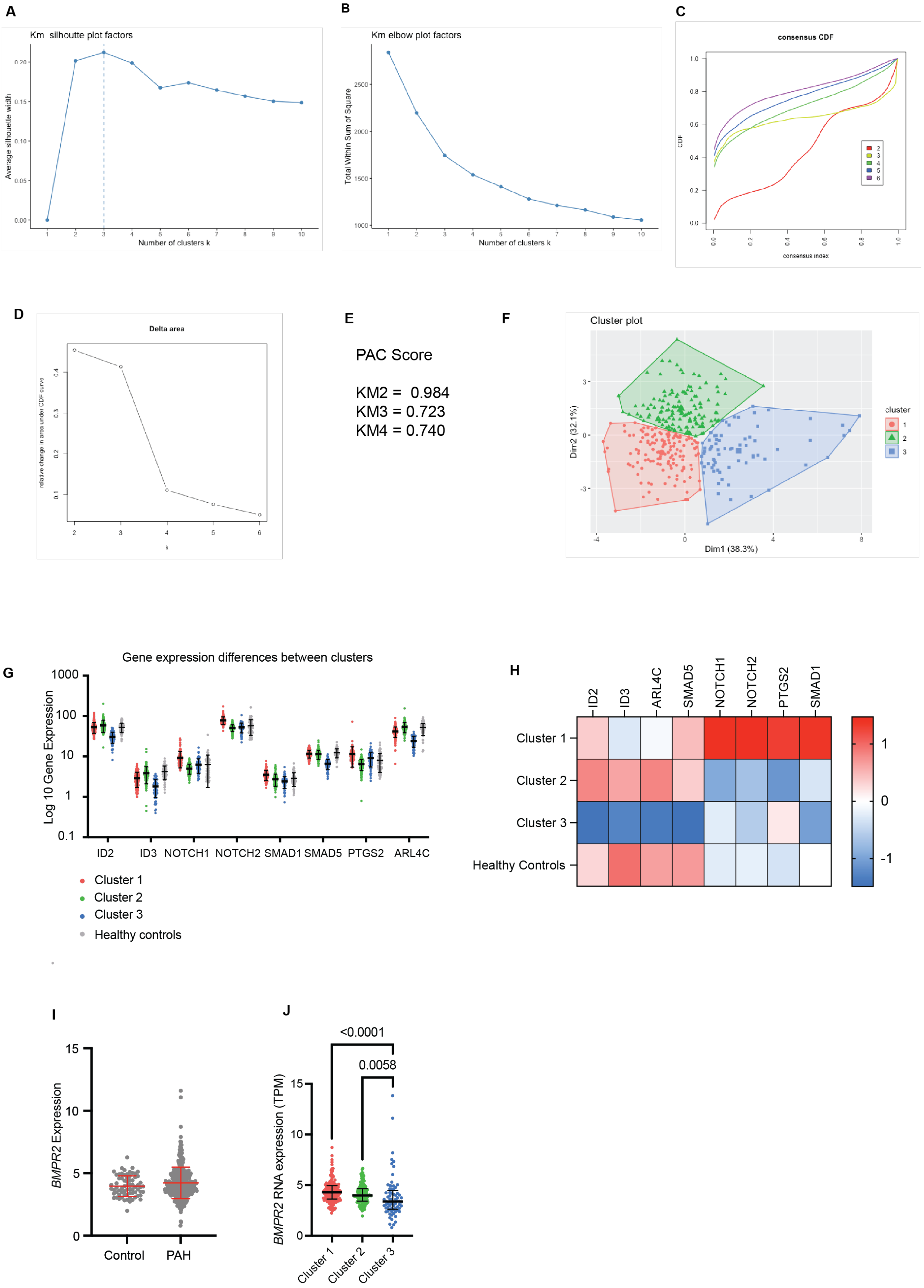
A
Km silhoutte plot factors
Average silhouette width
Number of clusters k
B
Km elbow plot factors
Total Within Sum of Square
Number of clusters k
C
consensus CDF
CDF
consensus index
D
Delta area
relative change in area under CDF curve
k
E
PAC Score
KM2 = 0.984
KM3 = 0.723
KM4 = 0.740
F
Cluster plot
Dim2 (32.1%)
Dim1 (38.3%)
cluster
G
Gene expression differences between clusters
Log 10 Gene Expression
ID2
ID3
NOTCH1
NOTCH2
SMAD1
SMAD5
PTGS2
ARL4C
Cluster 1
Cluster 2
Cluster 3
Healthy controls
H
ID2
ID3
ARL4C
SMAD5
NOTCH1
NOTCH2
PTGS2
SMAD1
Cluster 1
Cluster 2
Cluster 3
Healthy Controls
I
BMPR2 Expression
Control
PAH
J
<0.0001
0.0058
BMPR2 RNA expression (TPM)
Cluster 1
Cluster 2
Cluster 3

K

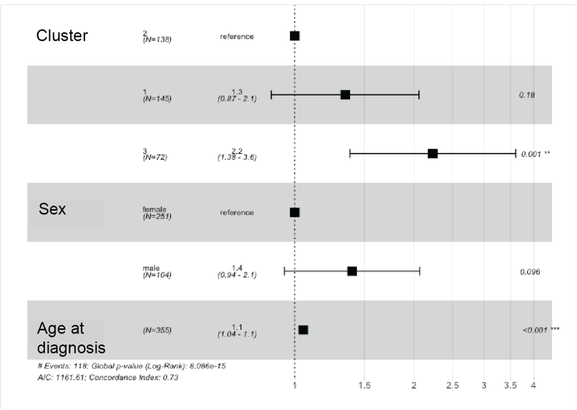


L

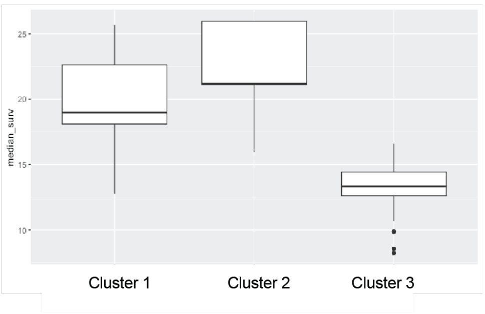


**Fig. S3. Supporting results for unsupervised machine learning clustering of the BMPR-II biomarker panel using RNAseq data from the UK PAH cohort study.** Justification for use of clustering algorithm Kmeans with K=3 with RNAseq data **(A)** Silhouette plot for Kmeans, **(B)** Elbow plot for Kmeans, **(C)** consensus CDF plot for Kmeans, **(D)** CDF delta area plot for Kmeans and **(E)** PAC scores for Kmeans=2-4, **(F)** Cluster plot for Kmeans = 3. **(G and H)** Log10 gene expression values of biomarker panel genes measured using RNAseq grouped according to consensus cluster (clusters 1-3), also showing healthy controls (grey) as a comparator group (N=67). Data shown as mean ± SD and as heatmap using Z-score. **(I)** *BMPR2* transcript expression (TPM) in control (N= 67) and PAH (N= 356). **(J)** *BMPR2* transcript levels (TPM) in patients stratified by cluster. Cluster 1 N=146, cluster 2 N=138, cluster 3 N=72).Data shown as median ± IQR. Kruskal Wallis statistical analysis between clusters. **(K)** Cox proportional hazard model between clusters including age at diagnosis and sex. **(L)** Boot strap resampling of survival data based upon Kmeans K=3 clustering analysis of RNAseq data. Repeat random sampling of the data set (N=355) was performed for 1000 bootstrap replicates and median survival ± IQR reported for each cluster. Kruskal Wallis statistical analysis for difference in median survival between clusters, p = < 0.00001.

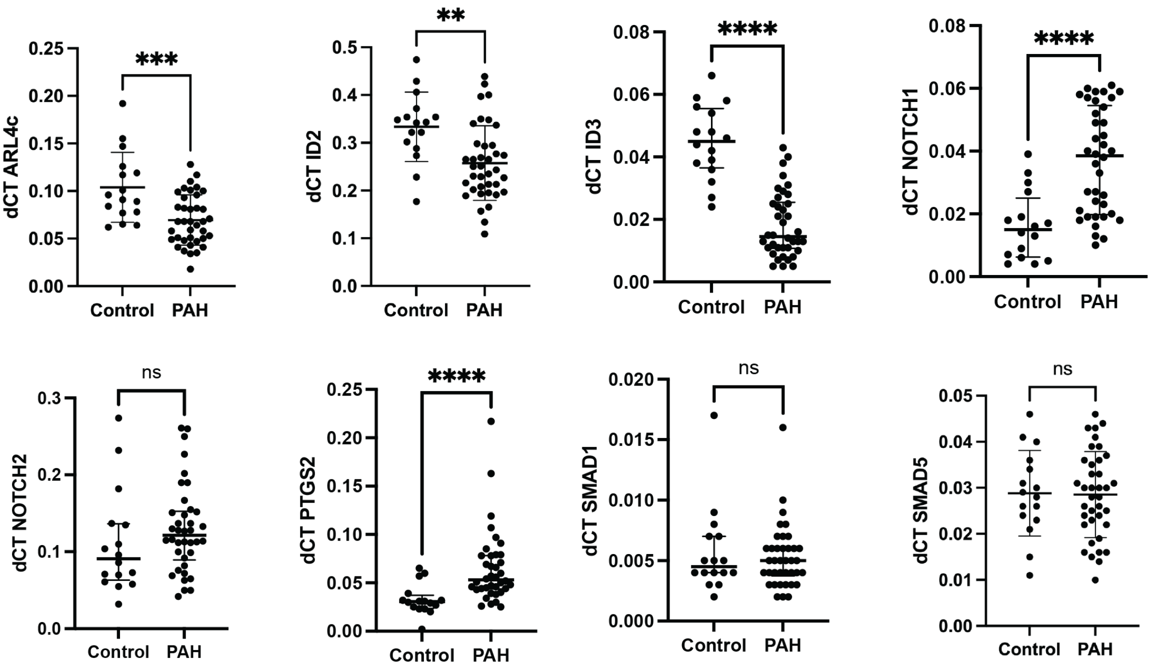


**Fig. S4. Validation cohort for the BMPR-II biomarker panel.** PAH patient samples from an international validation cohort of PAH (N=37) and healthy controls (N=16) were assessed for expression of our BMPR2 biomarker panel genes using qPCR. ΔCT values of BMPR2 biomarker panel genes normalised to 4 housekeeping genes. Data shown as mean ± SD or median ± IQR and statistics performed as students t-test or Mann-Whitney dependent upon data normality.

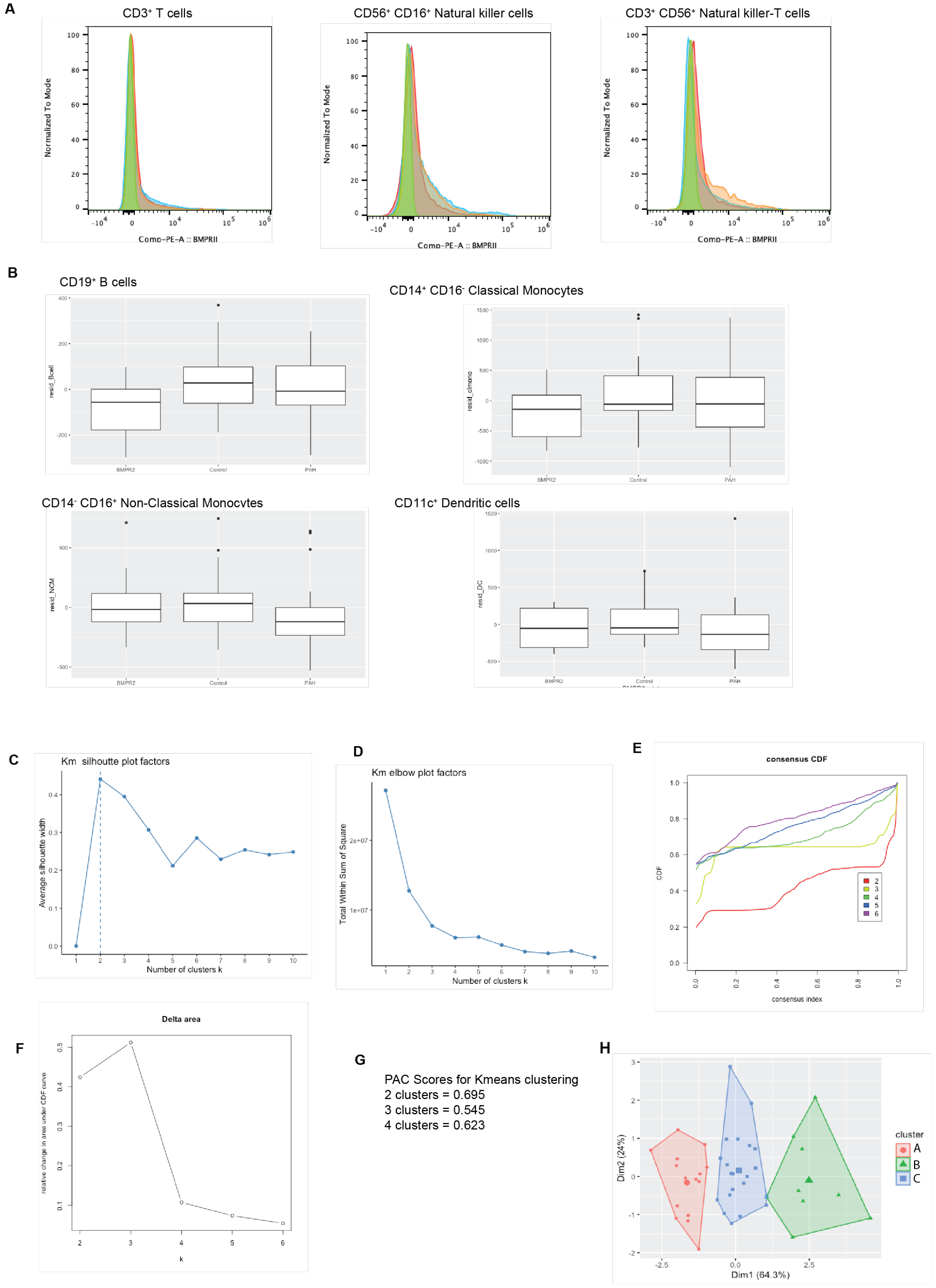
A
CD3+ T cells
CD56+ CD16+ Natural killer cells
CD3+ CD56+ Natural killer-T cells
Normalized To Mode
Comp-PE-A :: BMPRII
B
CD19+ B cells
resid_Bcell
CD14+ CD16- Classical Monocytes
resid_clmono
CD14- CD16+ Non-Classical Monocytes
resid_NCM
CD11c+ Dendritic cells
resid_DC
BMPR2
Control
PAH
C
Km silhoutte plot factors
Average silhouette width
Number of clusters k
D
Km elbow plot factors
Total Within Sum of Square
Number of clusters k
E
consensus CDF
CDF
consensus index
F
Delta area
relative change in area under CDF curve
k
G
PAC Scores for Kmeans clustering
2 clusters = 0.695
3 clusters = 0.545
4 clusters = 0.623
H
Dim2 (24%)
Dim1 (64.3%)
cluster
A
B
C

**I**

| Cluster A | Cluster B | Cluster C |
|---|---|---|
| R321X | R321X | E845X |
| BMPR2 C123R; SMAD9 R294X | | Deletion exon 1 |
| W9X | | Q850X |
| R332X | | |

**J**

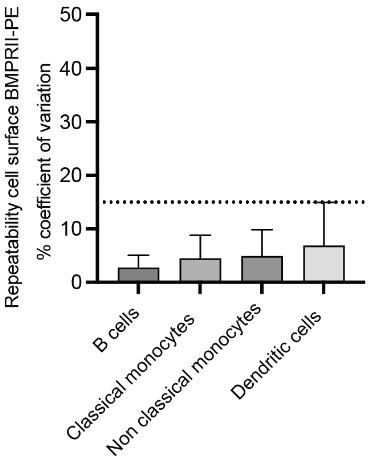


**K**

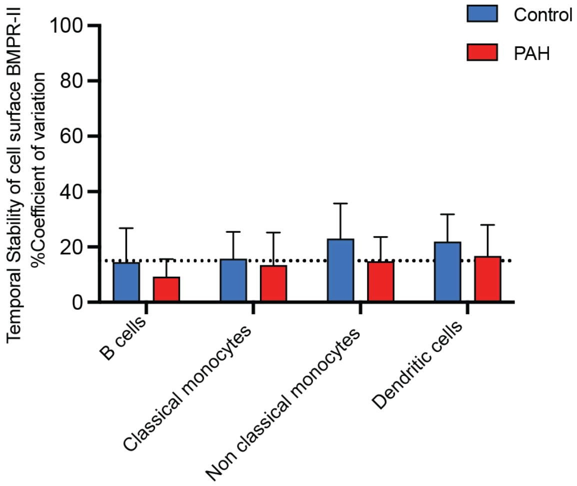


**Fig. S5. Supporting results for BMPR-II cell surface staining in leukocyte populations using samples from StratosPHere 1.** (A) BMPR-II cell surface staining in CD3+ T cells, CD56+ CD16+ natural killer cells, CD3+ CD56+ Natural killer T cells. Data shown as fluorescence of 3 representative healthy controls (red, blue, orange) versus an unstained control (green). **(B)** Cell surface BMPR-II levels in control (N=17), IPAH patient (N=20) and *BMPR2* mutation carriers (BMPR2) PAH (N=8) leukocyte populations after correction for BMPR2 antibody batch variation. Data shown as median ± IQR. Justification for use of clustering algorithm Kmeans with K=3 **(C)** Silhouette plot for Kmeans, **(D)** Elbow plot for Kmeans, **(E)** consensus CDF plot for Kmeans, **(F)** CDF delta area plot for Kmeans and **(G)** PAC scores for K=2-4. **(H)** Clusters of patients and controls using cell surface BMPR-II staining data using algorithm Kmeans K=3. **(I)** Distribution of *BMPR2*-PAH mutation carriers between cell surface BMPR2 clusters. **(J)** Technical repeatability of cell surface BMPR2 staining of duplicate samples was tested using co-efficient of variation in N=69 samples in B-cells, classical and non-classical monocytes and dendritic cells using both control and PAH patients. MFI values were compared. Data shown as mean %CV ± SD. Dotted line at 15%CV for assay assessment purposes. **(K)** Temporal variation of PAH patients (N=15) and healthy controls (N=12) between baseline and 16 weeks (± 4 weeks) in B-cells, classical and non-classical monocytes and dendritic cells tested using co-efficient of variation. Data shown as mean %CV ± SD. Dotted line at 15%CV for assay assessment purposes.

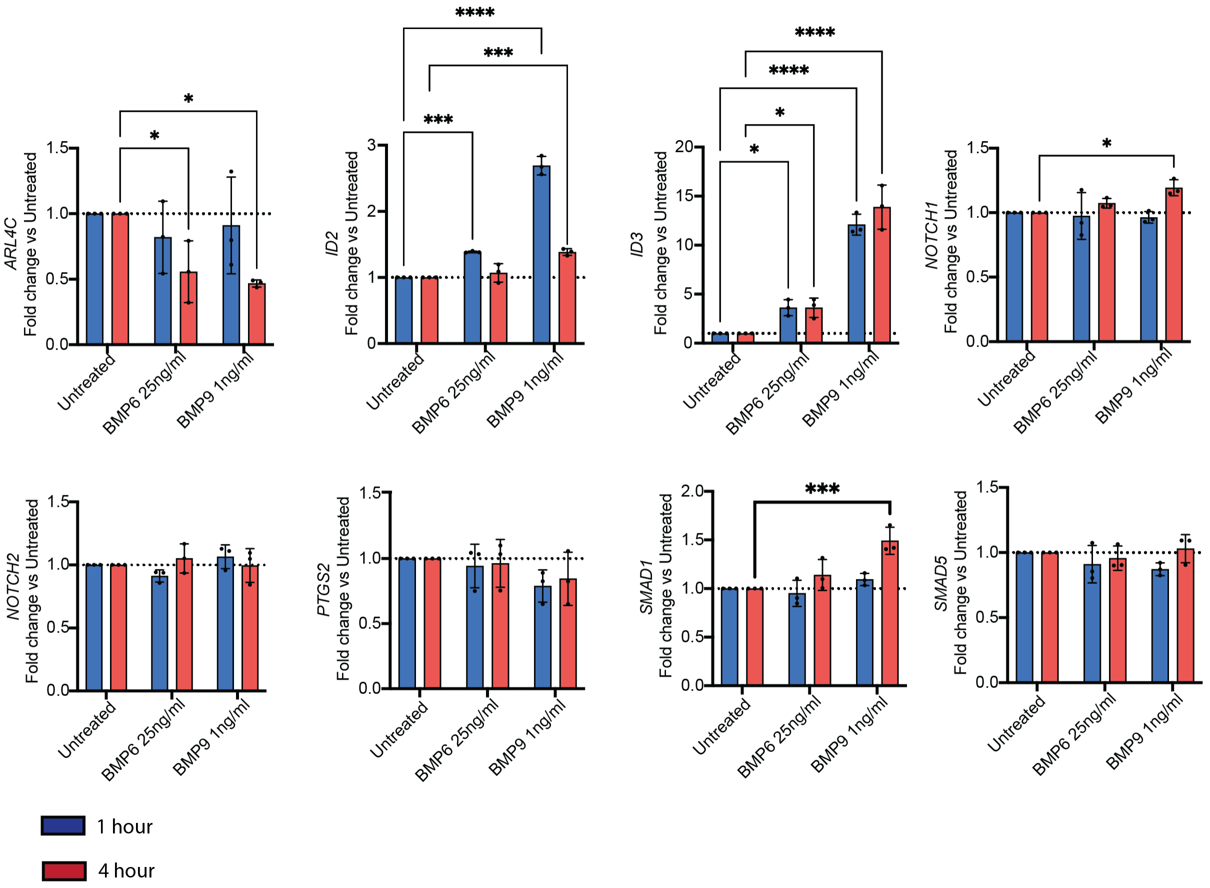


**Fig. S6. THP1 monocytic cell line treated with BMP ligands**.

THP1 cells were treated with 25ng/ml BMP6 or 1ng/ml BMP9 for 1 hour (blue) or 4 hours (red). Cells were harvested and analysed for change in expression of the BMPR-II biomarker panel genes. Data shown as mean fold change from the relevant time point untreated sample ± standard deviation for 3 technical replicates. Statistics shown as ANOVA compared to the untreated.

**Supplemental references**

44. G. Monaco, B. Lee, W. Xu, S. Mustafah, Y. Y. Hwang, C. Carré, N. Burdin, L. Visan, M. Ceccarelli, M. Poidinger, A. Zippelius, J. Pedro de Magalhães, A. Larbi, RNA-Seq Signatures Normalized by mRNA Abundance Allow Absolute Deconvolution of Human Immune Cell Types. *Cell Reports* **26**, 1627-1640.e7 (2019).
